# Developmental trajectories of aerobic endurance performance in highly trained youth soccer players aged 14 to 19 years

**DOI:** 10.64898/2025.12.04.25341632

**Authors:** Ludwig Ruf, Sascha Härtel, Stefan Altmann

**Author notes:** **Corresponding author:** Ludwig Ruf, Horrenberger Straße 58, 74939 Zuzenhausen, Germany.

## Abstract

**Purpose:** We aimed to identify distinct clusters of developmental trajectories in aerobic endurance performance among highly trained youth soccer players aged 14 to 19 years, moving beyond traditional group-level analyses to characterize potential heterogeneity of individual developmental pathways.

**Methods:** Mixed-longitudinal data from 149 male youth soccer players from a professional German academy were analyzed. Aerobic endurance was assessed using velocity at individual anaerobic threshold (vIANT) and velocity at 4 mmol.L^−1^ blood lactate (v4) from incremental treadmill tests. A hybrid clustering framework combining Generalized Additive Models (GAM) with model-based clustering (Mclust) was employed. Individual trajectories were estimated using GAMs with factor smooth basis functions, followed by comprehensive feature extraction (15 trajectory characteristics). Growth mixture models were applied across 3–7 clusters with multiple covariance structures, with model selection based on weighted consensus scoring emphasizing BIC, silhouette coefficient, and AIC.

**Results:** Four distinct developmental trajectory clusters were identified with consistent patterns across both vIANT and v4. The overall population mean trajectory demonstrated minimal systematic change across the age range. However, substantial between-player heterogeneity was evident: approximately one-fifth of players showed substantial improvement followed by stabilization, one-quarter exhibited progressive decline, while the majority displayed minimal change patterns with either slight improvement or slight decline. Endurance performance converged with increasing age across all clusters.

**Conclusion:** Aerobic endurance development in highly trained youth soccer players follows distinct individual trajectories rather than uniform patterns during adolescence. These findings highlight that group-level analyses mask meaningful individual differences, emphasizing the importance of individualized talent evaluation and training prescription.

## Introduction

Monitoring developmental trajectories of physical performance of youth soccer players plays an important role to optimally evaluate a player’s current and potential future physical performance and ultimately inform talent selection decisions (i.e., progression to the next age group within a developmental program). Recent reviews have emphasised the need for longitudinal studies within the realm of applied talent research which advances our understanding of how youth soccer players naturally develop during adolescence ^1,2^. The development of physical performance rarely follows a linear pattern from early adolescence (i.e., 12 years of age) through to adulthood (i.e., 20 years of age) and is influenced by a multitude of factors such as training history and biological maturation that drives physiological and morphological changes ^3^. Without understanding these individual trajectories, selection and evaluation processes risk misinterpreting temporary performance differences as stable characteristics. There is therefore a pressing need to characterize the individual developmental trajectories that underlie physical performance development in youth soccer players.

Endurance performance is one of several critical components of physical performance in soccer and is associated with match running output in youth soccer players ^4^. Several longitudinal observational studies have examined the development of physical performance, including endurance performance, in youth soccer players, but these investigations have predominantly focused on average developmental trajectories ^5^ or binary comparisons between selected vs. de-selected ^6^ or future professional vs. non-professional players ^7^. While these studies provide valuable insights into general developmental patterns, such broad classifications may obscure the heterogeneity of individual developmental pathways that exist during adolescence. At the group level, endurance performance, predominantly assessed using field-based shuttle running tests, has been shown to develop throughout the entire adolescent period, with longitudinal studies demonstrating consistent increases across all chronological ages and biological maturation stages ^5,7,8^. However, despite these well-established findings at the group level, empirical data regarding individual developmental trajectories in youth soccer players remain limited. Moreover, laboratory-based assessments of isolated aerobic capacity (e.g., lactate threshold measures) may reveal different developmental patterns compared to field-based shuttle tests that encompass multiple physiological and neuromuscular components. To advance talent evaluation and selection practices, there is a need to move towards identifying clusters of developmental trajectories in endurance performance that capture the true diversity of development in highly trained youth soccer players. This approach would provide more nuanced insights into the prevalence and magnitude of endurance development during adolescence.

Therefore, the aim of the current study was to identify clusters of developmental trajectories of aerobic endurance performance in highly trained youth soccer players aged 14 to 19 years.

## Methods

### Participants

The sample included aerobic endurance performance data available for N = 149 male academy soccer players and n = 1380 datapoints, respectively (chronological age range: 14.0 to 19.0 years), over a nine-year period (summer 2016 to summer 2025). Players belonged to the U14, U15, U16, U17, and U19 age group of one professional German youth academy and can be classified as tier 3 athletes (highly trained/national Level) competing at the highest national level within their respective age group competitions ^9^. Upon enrolment, parents/guardians signed contracts providing consent confirming that data arising as a condition of regular player monitoring procedures can be used for research purposes. Ethical approval was granted by the ethics committee of [blinded for peer-review] (ref.no. 24-10).

We acknowledge that no formal sample size calculation was performed for this study. While sample size calculation for clustering approaches has traditionally been challenging due to the exploratory nature of these methods, recent research has provided more systematic guidance for these analyses ^10^. Sample size requirements for Mixture Models depend on multiple factors including the degree of separation between latent classes, the relative size of the smallest clusters, the number of measured variables, and the specific clustering method employed ^11^. Given the exploratory nature of our functional mixture modelling approach, we therefore employed a comprehensive model diagnostic process to ensure the validity and interpretability of the clustering solution, including a comparison with a traditional parametric growth mixture modelling approach.

### Study design

Retrospective mixed-longitudinal observational study design.

### Procedures

Players performed indoor an incremental test on a Woodway treadmill (Woodway GmbH, Weil am Rhein, Germany) to determine lactate-based thresholds ^6,12^. The slope of the treadmill was set at 1% incline. Running velocity started at 6 kmh-1 and increased by 2 kmh-1 with each stage lasting 3 min. Between stages, a 30-second passive recovery phase was implemented allowing for capillary blood collection from the earlobe. Players received instructions to perform the test until exhaustion. Blood lactate concentration analysis for each stage was performed with the Biosen C-Line Sport system (EKF-diagnostics GmbH, Barleben, Germany). The Ergonizer Software (K. Roecker, Freiburg, Germany) was used to compute the following outcome measures: i) velocity at the individual anaerobic threshold (velocity at which blood lactate concentration was 1.5 mmol.L^−1^ above the individual aerobic threshold, determined as the velocity at which blood lactate concentration begins to rise above baseline levels, vIANT) and ii) velocity at the absolute lactate concentration of 4 mmol.L^−1^ (v4). We acknowledge that several other threshold concepts exist and aimed to describe two fundamentally different ones here: an absolute threshold based on a fixed blood lactate concentration (v4) and an individual threshold accounting for the athlete’s personal lactate kinetics (vIANT) ^13^.

### Statistical analyses

Statistical analyses were conducted in RStudio (version 1.2.5033). A detailed description of the entire clustering methodology can be found in the Supplementary File (Supplementary 1. Comprehensive clustering methodology).

#### Data preparation

Player inclusion required adequate longitudinal data coverage whereby players required data spanning at least 3 distinct ages, a minimum 2-year temporal span, and at least 3 observations. We implemented a hybrid-based clustering framework combining Generalized Additive Models with model-based clustering (GAM + Mclust), with Latent Class Mixed Models (LCMM) computed as a comparative methodological approach detailed in the Supplementary File (Supplementary 2. Results from the Latent Class Mixed Models (LCMM) clustering approach).

#### Step 1: Individual Trajectory Estimation

Generalized Additive Models were implemented using the mgcv package (version 1.9-1) with factor smooth basis functions to estimate player-specific developmental trajectories for both outcome measures vIANT and v4 across chronological age. Optimal basis dimensions (k = 3-6) were determined through multi-criteria evaluation incorporating deviance explained, information criteria (BIC, AIC), and effective degrees of freedom. Based on this systematic assessment, k = 3 was selected as optimal for vIANT and k = 4 for v4, representing the best balance between model fit and computational stability.

#### Step 2: Feature-Based Growth Mixture Modelling

Individual predicted trajectories were systematically generated across age intervals from 14 to 19 years with 0.1-year increments. Comprehensive feature extraction yielded 15 trajectory characteristics including endpoints, developmental rates, curvature indices, and variability metrics. All features were standardized to ensure equal contribution to the clustering process. Growth mixture models were computed using the mclust package (version 6.1.1) across 3-7 clusters and 8 covariance structures representing different assumptions about cluster geometry. Model selection employed weighted consensus scoring emphasizing BIC (30%), silhouette coefficient (30%), and AIC (25%) over traditional distance-based metrics (Calinski-Harabasz: 10%, Dunn: 2%, Davies-Bouldin: 3%), reflecting the importance of both statistical model adequacy and meaningful cluster distinction. We identified a 5-cluster solution with spherical, equal-volume covariance structure as demonstrating optimal performance.

Individual predicted trajectories were systematically generated across age intervals from 14 to 19 years with 0.1-year increments. Comprehensive feature extraction yielded 15 trajectory characteristics including performance levels, developmental rates, variability metrics, timing features, and curvature indices. All features were standardized to ensure equal contribution to the clustering process. Model-based clustering was performed using the mclust package (version 6.1.1) across 3-7 clusters and multiple covariance structures (i.e., EVI, VEI, VVI). Model selection employed weighted consensus scoring emphasizing BIC (30%), silhouette coefficient (30%), and AIC (25%) over traditional distance-based metrics (Calinski-Harabasz: 10%, Dunn: 2%, Davies-Bouldin: 3%). The optimal solution for both vIANT and v4 identified 4 clusters with the VEI covariance structure.

#### Model Diagnostics

GAM model validation included residual analysis for homoscedasticity assessment, observed vs. predicted value evaluation, and person-level model fit quality distribution. Mclust clustering validation encompassed posterior probability analysis for assignment certainty, silhouette analysis for cluster separation quality, bootstrap stability assessment using Adjusted Rand Index across 30 -50 resampling iterations, distance-based validation metrics, entropy-based separation analysis, and systematic outlier detection using Mahalanobis distances. A comprehensive overview of the clustering model diagnostic results can be found in the Supplementary File (Supplementary 3. Comprehensive model diagnostics for vIANT of both clustering approaches (Hybrid approach (GAM + Mclust) and Latent Class Mixed Model (LCMM)) and Supplementary 4. Comprehensive model diagnostics for v4 of both clustering approaches (Hybrid approach (GAM + Mclust) and Latent Class Mixed Model (LCMM))).

##### Reference overall mean trajectory estimation

To estimate the overall mean trajectory of the entire sample, we fitted again a generalized additive model using the mgcv package (version 1.9-1) with restricted maximum likelihood (REML). Chronological age served as the independent variable and both outcome measures vIANT and v4 as the dependent variable. Random intercepts for player ID were modelled as ridge-penalized smooths to account for repeated measures within individuals. Optimal basis functions (tp, cr, cs, ps) and smoothness parameters (k = 3 to 6) were determined by minimising the Akaike Information Criterion (AIC). Population-level predictions were obtained by excluding the random effect term, providing the marginal mean trajectory with correct confidence intervals. Detailed model information is provided in the Supplementary File (Supplementary 5. Comparative quality assessment across both outcome measures).

## Results

Four distinct clusters of developmental trajectories were identified for both outcome measures using the two-step hybrid framework combining individual GAM trajectory estimation followed by feature-based clustering. Figures 1 and 2 illustrate the individual and GAM model-predicted cluster mean developmental trajectories together with the mean trajectory of the entire sample as reference for vIANT and v4, respectively.

**Figure 1.**
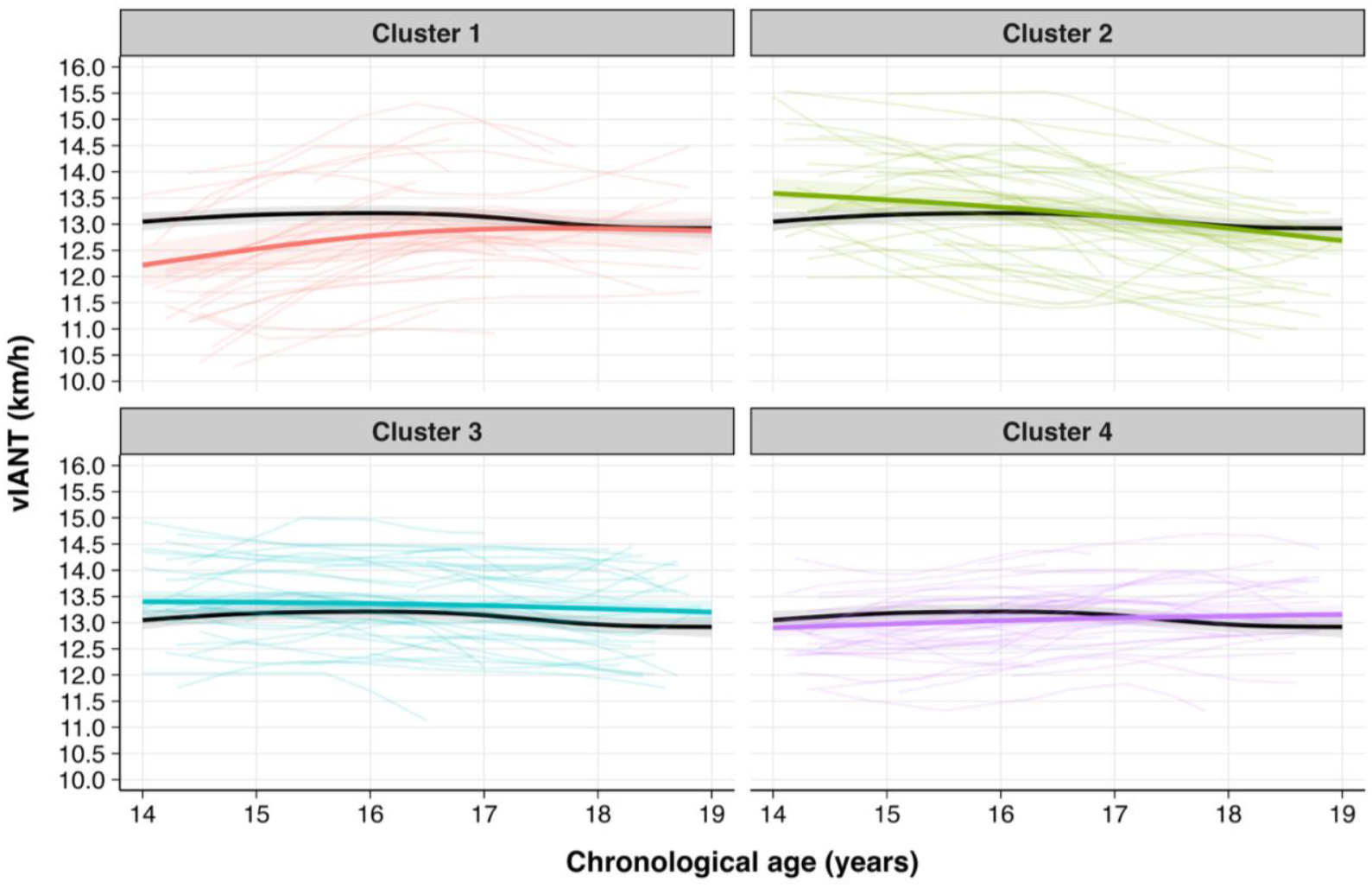
Distinct trajectory clusters for the outcome measure *velocity at the individual anaerobic threshold (vIANT)* derived from the two-step hybrid approach combining individual GAM trajectory estimation followed by feature-based clustering. Each cluster shows individual trajectories and GAM model-predicted cluster means with model-based 95% confidence intervals. The black line represents the mean trajectory of the entire sample as reference with 95% confidence intervals.

**Figure 2.**
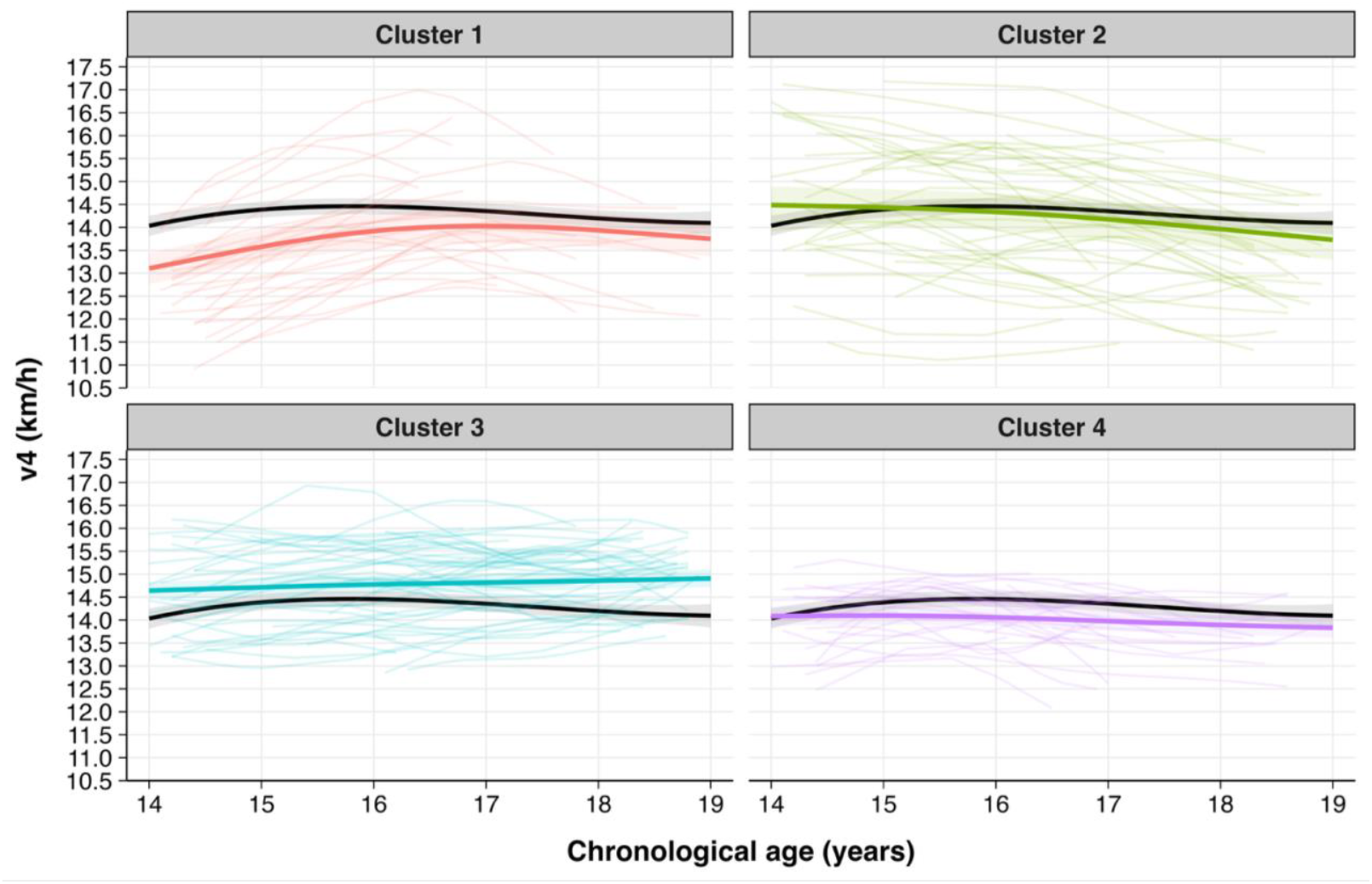
Distinct trajectory clusters for the outcome measure *velocity at lactate concentration of 4 mmol.L^−1^ (v4)* derived from the two-step hybrid approach combining individual GAM trajectory estimation followed by feature-based clustering, showing individual trajectories and GAM model-predicted cluster means with model-based 95% confidence intervals. The black line represents the mean trajectory of the entire sample as reference with 95% confidence intervals.

For vIANS, the clusters can be characterised as: substantial improvement then stabilization (cluster 1, n = 26, 17%), progressive decline (cluster 2, n = 36, 24%), minimal decline (cluster 3, n = 45, 30%), and minimal improvement (cluster 4, n = 42, 28%) trajectories.

For v4, similar trajectory patterns emerged: substantial improvement then stabilization (cluster 1, n = 26, 17%), progressive decline (cluster 2, n = 36, 24%), minimal improvement (cluster 3, n = 58, 39%), and minimal decline (cluster 4, n = 29, 19%) trajectories.

## Discussion

In this study, we identified distinct clusters of developmental trajectories of aerobic endurance performance using mixed-longitudinal data from highly trained youth soccer players aged 14 to 19 years. The overall mean trajectory demonstrated minimal systematic change across the age range, with aerobic endurance performance remaining relatively stable between 14 and 19 years. However, this average pattern masks considerable individual variation. Across both outcome measures vIANT and v4, approximately one-fifth of players demonstrated substantial improvement followed by stabilisation, one-quarter showed progressive decline, while the majority exhibited minimal change patterns with either slight improvement or slight decline. These findings provide exploratory evidence for the heterogeneity of individual developmental trajectories and challenge the assumption of consistent improvement of aerobic endurance performance during adolescence.

The heterogeneous trajectory patterns likely reflect the complex interplay of training history, accumulated training load, and individual factors during adolescence ^3^. We observed similar cluster patterns between vIANT and v4 which can be explained by their strong correlation (repeated measures correlation = 0.85, 95% CI: 0.83-0.86), highlighting that both outcome measures represent the same construct of aerobic endurance performance. We observed four distinct trajectory patterns which may be explained by multiple developmental factors during adolescence, including asynchronous development of aerobic and anaerobic endurance ^3^, varying training histories and accumulated loads, and systematic shifts in training emphasis toward anaerobic power and neuromuscular qualities as players approach maturity. For example, cluster 1 (“substantial improvement then stabilisation”) likely reflects predominant aerobic development until 16-17 years, followed by physiological transition toward anaerobic development ^14^. Conversely, cluster 2 (“progressive decline”) challenges previous observations of continuous development of endurance performance during adolescence ^3^, potentially indicating prioritisation shifts toward anaerobic and neuromuscular adaptations at the expense of aerobic endurance ^15^.

Despite the stable overall mean trajectory, substantial between-player heterogeneity highlights that group-level analyses mask meaningful individual differences in aerobic endurance development. The convergence of performance levels with age (i.e., narrowing the spread of cluster means) indicates that the accumulation of systematic training reduces initial disparities. Players with superior aerobic endurance may accept slight declines while potentially focusing on other physical qualities (e.g., speed and power), whereas players with poorer aerobic endurance performance improve toward sufficient levels. These findings underscore the importance of focusing on individual developmental trajectories rather than average development curves for talent evaluation and training prescription. Previous research has identified conflicting evidence for potential critical periods in aerobic endurance development (12-14 vs 14-16 years) ^16^, and recent longitudinal studies emphasise individual variation in both developmental timing and trainability of aerobic fitness ^14^, supporting our findings of distinct developmental trajectories rather than universal critical periods.

Our two-step hybrid framework advances beyond traditional trajectory approaches ^5^ by capturing individual heterogeneity that group-level classifications ^6,7^ typically miss. The GAM modelling approach accounts for non-linear developmental patterns, addressing limitations of studies focusing solely on group-level trends ^8^. Our findings contrast with previous literature reporting consistent increases in endurance performance throughout adolescence ^5,7,8^. This discrepancy may be attributed to test-specificity: our laboratory-based aerobic threshold measures (vIANT, v4) isolate aerobic capacity, whereas commonly used field-based tests such as the Yo-Yo Intermittent Recovery Test assess a broader construct encompassing aerobic, anaerobic, and neuromuscular qualities ^13,17^. This methodological difference may explain why we observed minimal average development in pure aerobic capacity whilst studies that included endurance performance tests encompassing multidimensional constructs demonstrated improvements throughout adolescence. This aligns with calls for longitudinal talent research ^1,2^ and offers practitioners a more nuanced understanding of individual developmental trajectories.

### Limitations

Despite the novelty and practical applicability of our trajectory clustering approach for understanding aerobic endurance development, several limitations should be acknowledged. First, we did not assess biological maturation status, though previous research suggests minimal associations between skeletal maturation and aerobic performance in youth soccer players ^18^. Second, our findings are derived from a single professional academy, which may limit generalizability to other contexts and playing levels. Third, the relatively small sample sizes within some clusters require cautious interpretation of cluster-specific patterns. Fourth, our analysis was restricted to chronological ages 14 to 19 years, not encompassing the entire adolescent period and adulthood. Finally, we focused exclusively on aerobic endurance without considering other physical performance qualities (e.g., strength, speed, power), which prevents holistic player profiling that might further explain the observed developmental trajectories. Future research should address these limitations through multi-site studies incorporating early adolescence and adulthood, larger samples, and comprehensive physical performance assessments.

## Practical Applications

Our findings have immediate practical implications for talent development programs in youth soccer. To translate these findings into practice, we developed an interactive web-based application (https://rufludwig.shinyapps.io/endurance-clustering/) that allows practitioners to input longitudinal aerobic endurance performance data (minimum 3 data points of a player) to receive cluster membership probabilities with trajectory visualisations. Understanding a player’s individual developmental trajectory enables practitioners to make better informed evaluations of current and potential future aerobic endurance performance, ultimately supporting talent selection decisions.

## Conclusion

This study demonstrates that aerobic endurance development in highly-trained youth soccer players follow distinct individual trajectories rather than uniform patterns during adolescence. While the overall mean trajectory remained relatively stable between 14 and 19 years, substantial between-player heterogeneity was evident, with approximately one-fifth demonstrating substantial improvement, one-quarter showing progressive decline, and the majority exhibiting minimal change patterns. These results offer nuanced insights for more individualized talent evaluation and selection practices within highly-trained youth soccer players.

## Supporting information

2025_11_28_supplementary_file_medRxiv

## Data Availability

All data produced in the present study are available upon reasonable request to the authors

## Acknowledgements

We would like to thank the Department of Performance Diagnostics of the TSG Hoffenheim for their support during data collection across the past years.

