## Supplementary material for "Developmental trajectories of aerobic endurance performance in highly trained youth soccer players aged 14 to 19 years": 2025_11_28_supplementary_file_medRxiv

**Journal:** Preprint Server medRxiv

**Article Type:** Brief Report

### Glossar

This glossary defines all model selection and clustering quality metrics reported in Supplementary 1 to 5. For interpretation thresholds and comprehensive clustering methodology, see Supplementary 1.

#### *Model selection criteria*

| Metric | Definition | Construct Measured | Interpretation |
|---|---|---|---|
| **Log-likelihood (loglik)** | Logarithm of the likelihood function evaluated at estimated parameters | Overall model fit | Higher (less negative) values indicate better fit to observed data |
| **Akaike Information Criterion (AIC)** | Information-theoretic criterion: $AIC = 2k - 2\log(L)$ | Parsimony-adjusted model fit | Lower values indicate better balance between fit and complexity |
| **Bayesian Information Criterion (BIC)** | Similar to AIC with stronger complexity penalty | Model comparison and parsimony | Lower values indicate better model balancing fit and simplicity |
| **Deviance Explained / $R^2$** | Proportion of variability explained by model predictors | Explained variance | Higher values indicate more captured variance (range: 0 to 1) |
| **Effective Degrees of Freedom (edf)** | Estimated parameters controlling smoothness in GAMs | Model flexibility and complexity | Higher values indicate more flexible smooth terms |

#### *Cluster quality metrics*

| Metric | Definition | Construct Measured | Interpretation |
|---|---|---|---|
| **Silhouette Width** | $s(i) = (b(i) - a(i)) / \max(a(i), b(i))$ where $a(i)$ = mean within-cluster distance, $b(i)$ = minimum mean distance to other clusters | Cluster compactness and separation | Near 1 = excellent separation; near 0 = overlap; negative = potential misclassification |
| **Dunn Index** | Ratio of minimum inter-cluster distance to maximum intra-cluster distance | Cluster separation relative to compactness | Higher values indicate more distinct and internally cohesive clusters |
| **Davies-Bouldin Index (DB)** | Average ratio of within-cluster scatter to between-cluster separation | Cluster distinctness and overlap | Lower values indicate more compact and well-separated clusters |

| Metric | Definition | Construct Measured | Interpretation |
|---|---|---|---|
| **Calinski-Harabasz Index (CH)** | Ratio of between-cluster dispersion to within-cluster dispersion | Cluster cohesion and separation | Higher values indicate better defined cluster structure |
| **Entropy** | Information-theoretic measure of uncertainty in cluster assignments | Classification certainty | Lower values indicate more confident and unambiguous assignments (range: 0 to 1) |
| **Mean Classification Uncertainty** | Average uncertainty across individuals: 1 − maximum posterior probability | Assignment confidence | Lower values indicate more precise cluster allocations |
| **Bootstrap Stability (Mean ARI)** | Mean Adjusted Rand Index across bootstrap resamples | Cluster stability and reproducibility | Higher values indicate consistent cluster recovery across samples (range: 0 to 1) |
| **Bootstrap Stability (SD)** | Standard deviation of ARI values across resamples | Consistency of stability | Lower values indicate uniform stability across bootstrap samples |

### Supplementary 1. Comprehensive clustering methodology

#### *Workflow Approach*

We implemented a two-step hybrid analytical framework (individual trajectory estimation followed by feature-based Growth Mixture Modelling, Hybrid approach; GAM + Mclust) to identify distinct developmental trajectory clusters, with conventional Growth Mixture Modelling computed as a comparative methodological approach. The hybrid approach was designed to leverage the flexibility of Generalized Additive Models (GAMs) for capturing individual developmental complexity while utilizing the probabilistic framework of mixture modelling for trajectory-based clustering.

#### *Data Preparation*

Player inclusion required adequate longitudinal data coverage whereby players required data spanning at least 3 distinct ages, a minimum 2-year temporal span, and at least 3 observations. These stringent criteria were established to ensure robust individual trajectory estimation: the requirement for at least 3 distinct integer ages (e.g., 13, 14, 15 years) ensured measurements across distinct chronological periods rather than multiple measurements within a single chronological age; the minimum 2-year temporal span (e.g., 13 to 15 years) guaranteed sufficient developmental coverage between first and last measurements; and the minimum 3 observations per individual provided adequate data points for reliable longitudinal modelling.

#### *Two-step Hybrid Framework: Individual Trajectory Estimation Followed by Feature-based Growth Mixture Modelling (Hybrid approach; GAM + Mclust)*

<u>Step 1: Individual Trajectory Estimation (GAM)</u>

Generalized Additive Models (GAMs) were implemented using the mgcv package (version 1.9-1) with factor smooth basis functions to estimate player-specific developmental trajectories for both outcome measures vIANT and v4 across chronological age. The factor smooth approach (bs="fs") creates separate smooth functions for each player ID while sharing smoothing parameters across individuals, effectively modelling individual-specific trajectories while maintaining statistical efficiency and preventing overfitting.

*Model Fitting Process*

Models were fitted using Restricted Maximum Likelihood (REML) estimation to optimize smoothing parameters while avoiding overfitting. The REML approach provides unbiased estimates of smoothing parameters and is particularly suitable for mixed-effects smoothing models with repeated measures data. The model structure accommodated both individual-specific developmental patterns and population-level age trends through the factor smooth framework.

Optimal basis dimensions (k = 3, 4, 5, 6, 7) were determined through multi-criteria evaluation incorporating deviance explained, information criteria (BIC, AIC), effective degrees of freedom, and convergence status. For each k-value, comprehensive performance metrics were extracted including: deviance explained (proportion of variance accounted for by the model), Akaike Information Criterion (AIC) and Bayesian Information Criterion (BIC) for model comparison and parsimony assessment, effective degrees of freedom (edf) as a model complexity measure, Generalized Cross-Validation (GCV) scores for predictive accuracy evaluation, REML likelihood values, and model convergence verification.

Rather than relying on single metrics, a composite scoring method was employed to balance model fit (deviance explained), parsimony (BIC penalty for complexity), predictive accuracy (AIC), and model complexity (edf). The combined score was calculated as: combined_score = scale_minmax(dev_explained) + scale_minmax(-BIC) + scale_minmax(-AIC) + scale_minmax(-edf), with all metrics normalized to [0,1] scales for equal contribution weighting.

*Model Selection Process*

Potential overfitting was systematically identified through: marginal improvements in $R^2$ beyond specific k-values (improvement <0.002 indicating diminishing returns), substantial BIC increases indicating excessive complexity penalties, and disproportionate increases in effective degrees of freedom relative to performance gains. Based on the combined scoring approach and overfitting evidence, optimal k-value was selected representing the best balance between model fit and complexity while avoiding overfitting observed at higher k-values.

*Model Diagnostics Process*

Comprehensive diagnostic evaluation was implemented through systematic diagnostic plots and performance assessments. Global performance metrics included deviance explained (proportion of total variance accounted for by the model), Generalized Cross-Validation (GCV) score as a measure of predictive accuracy with penalty for model complexity, and adjusted R-squared accounting for degrees of freedom consumption.

Residual analysis was conducted at both global and individual levels through multiple diagnostic visualizations: Residuals vs Fitted values plots for homoscedasticity assessment and detection of systematic patterns, Observed vs Predicted values plots for model accuracy evaluation with correlation analysis, Person-level fit quality distribution histograms showing RMSE per individual to identify systematic misfits, and Residuals vs Chronological Age plots for age-related bias detection.

Statistical assumption verification included homoscedasticity evaluation through Breusch-Pagan test approximation examining the relationship between squared residuals and fitted values, independence verification through examination of residual patterns across calendar age, and linearity assessment through residual versus fitted value plots to detect non-linear patterns indicating model misspecification.

Performance categorization was systematically applied with models classified as: Excellent (≥80% deviance explained), Good (70-79% deviance explained), Moderate (50-69% deviance explained), or Poor (<50% deviance explained), with corresponding recommendations for model improvement or acceptance.

### Step 2: Feature-Based Growth Mixture Modelling (Mclust)

Individual predicted trajectories were systematically generated from the optimal GAM model across age intervals from 14 to 19 years with 0.1-year increments, creating fine-grained individual-specific predicted developmental curves spanning the complete developmental period of interest. This approach provided high-resolution trajectory characterization essential for comprehensive feature extraction and subsequent clustering analysis.

#### *Feature Extraction Process*

Comprehensive feature extraction yielded 15 trajectory characteristics capturing distinct aspects of developmental patterns. Extracted features encompassed: trajectory endpoints (start_value at age 14, end_value at age 19), central tendency measures (mean_value, max_value, min_value across the chronological age span), overall developmental patterns (slope calculated as (end_value - start_value)/(age_range)), developmental phase analysis (early_slope for ages 14-15.6, late_slope for ages 16.3-19), variability metrics (volatility as standard deviation, range_value as maximum minus minimum value, max_change as maximum change between consecutive time points), temporal characteristics (age_at_max and age_at_min for chronological ages at maximum and minimum values), curvature assessment (curvature as mean of second differences quantifying trajectory non-linearity), and relative development measures (relative_change as proportional change from baseline).

All features were standardized using the scale() function to ensure equal contribution to the clustering process, converting all features to z-scores with mean = 0 and standard deviation = 1. This preprocessing step was critical for preventing features with larger numerical scales from dominating distance calculations and ensuring optimal mixture model performance across all trajectory characteristics.

*Model Selection Process*

Model selection employed a systematic evaluation approach across multiple dimensions: cluster numbers (G = 3 to 7) to balance interpretability with model complexity, and covariance structures representing different assumptions about cluster geometry including EVI (Equal volume, Variable shape, coordinate axes), VEI (Variable volume, Equal shape, coordinate axes), VVI (Variable volume, Variable shape, coordinate axes), and EEE (Equal volume, Equal shape, Equal orientation).

For each model combination (cluster number × covariance structure), comprehensive clustering validation metrics were computed including: model fit criteria (BIC - Bayesian Information Criterion for model comparison with complexity penalty, AIC - Akaike Information Criterion for predictive accuracy assessment), cluster separation assessment (Dunn index calculating the ratio of minimum inter-cluster distance to maximum intra-cluster distance), cluster compactness evaluation (Davies-Bouldin index measuring average similarity between clusters), cluster validity measurement (Calinski-Harabasz index computing the ratio of between-cluster variance to within-cluster variance), and individual assignment quality (silhouette coefficient calculating mean silhouette width across all observations).

To avoid over-reliance on single evaluation criteria, a weighted consensus approach was implemented prioritizing metrics appropriate for trajectory modelling. The weighting scheme emphasized statistical model adequacy and trajectory-specific clustering quality: BIC (30% weight) for model selection with complexity penalty, Silhouette coefficient (30% weight) for individual trajectory assignment quality, and AIC (25% weight) for predictive model fit. Traditional distance-based separation metrics received moderate to low weighting: Calinski-Harabasz Index (10% weight), Dunn Index (2% weight), and Davies-Bouldin Index (3% weight), reflecting that developmental trajectories naturally exhibit some overlap due to realistic individual developmental differences.

The optimal model was selected based on the highest composite score, balancing model fit, cluster separation, and parsimony across multiple criteria rather than relying on a single metric.

*Model Diagnostics Process*

Rigorous model validation was implemented through comprehensive diagnostic visualizations and summary reports encompassing multiple analytical perspectives. Cluster size distribution was assessed through bar plots showing number and percentage of observations per cluster to evaluate cluster balance and representativeness.

Posterior probability and assignment analysis included extraction of individual posterior probabilities for each player across all clusters from the fitted mixture model, with hard cluster assignments determined by maximum posterior probability class membership. Assignment uncertainty was systematically quantified as (1 - maximum posterior probability), providing measures of classification confidence. Box plots displayed posterior probability uncertainty by cluster with established quality classifications: high certainty (uncertainty ≤ 0.2), moderate certainty (0.2 < uncertainty ≤ 0.4), and low certainty (uncertainty > 0.4).

Distance-based validation metrics utilized Euclidean distance matrices computed from the standardized 15-dimensional feature space. Silhouette analysis was conducted calculating individual silhouette coefficients $s(i) = (b(i) - a(i)) / \max(a(i), b(i))$ where $a(i)$ represents mean intra-cluster distance and $b(i)$ represents mean distance to nearest neighboring cluster. Bar plots displayed cluster-specific average silhouette widths with established quality interpretation thresholds: ≥0.7 excellent separation, ≥0.5 good separation, ≥0.25 moderate separation, <0.25 poor separation.

Cluster separation and compactness indices were systematically displayed through bar plots showing multiple validation metrics with quality assessments. The Dunn Index was computed as the ratio of minimum between-cluster distance to maximum within-cluster distance, with quality benchmarks: ≥1.0 excellent separation, ≥0.5 adequate separation, <0.5 poor separation. The Davies-Bouldin Index measured average cluster similarity ratios relative to cluster separation, with quality thresholds: ≤1.0 excellent compactness, ≤2.0 good compactness, >2.0 poor compactness. The Calinski-Harabasz Index computed the ratio of between-cluster sum of squares to within-cluster sum of squares, with higher values indicating better-defined cluster separation.

Bootstrap stability assessment was conducted through 30 resampling iterations with 80% data subsampling. Models were refitted using identical parameters on bootstrap samples, and stability was quantified through Adjusted Rand Index (ARI) calculations comparing original and bootstrap cluster assignments. Histogram distributions showed ARI values across bootstrap iterations with established stability thresholds: ARI ≥0.8 excellent stability, ≥0.6 good stability, ≥0.4 moderate stability, <0.4 unstable solution.

Feature-space quality assessment was performed through box plots showing Mahalanobis distances by cluster with regularized covariance matrices to handle potential singularity issues in high-dimensional feature spaces. Chi-square distribution thresholds (97.5th percentile) were applied for outlier classification, enabling systematic identification of observations with atypical feature-space positioning relative to their assigned clusters.

Comprehensive summary reports integrated all diagnostic measures providing multi-dimensional quality assessments with overall solution ratings (excellent/good/moderate/poor) based on metric consensus and specific problematic areas identification for targeted improvement recommendations.

#### *Conventional Growth Mixture Modelling: Comparative Methodological Approach*

Traditional Growth Mixture Modeling (GMM) was examined using the lcmm package (version 2.1.0) as a direct clustering baseline for methodological comparison. Latent Class Mixed Models (LCMM) were systematically fitted with splines link functions specifically designed for bounded outcome variables, ensuring appropriate modelling of both outcome measures vIANT and v4 values.

##### Model Specification Process

Model selection employed a systematic grid search approach encompassing 15 model combinations (3 df values × 5 cluster numbers) to identify optimal model complexity and cluster number simultaneously. The model specification incorporated natural spline fixed effects structures (vIANT ~ ns(CalendarAge, df)) with varying degrees of freedom (df = 3, 4, 5) to capture potential non-linear developmental patterns across chronological age. Random intercepts (~ 1) were included to accommodate individual-specific baseline differences in outcome values.

Base models (ng = 1) were initially established as single-class models for each spline configuration providing stable starting values for subsequent mixture modelling procedures. Growth mixture models were then systematically evaluated across 3-7 latent clusters (ng = 3 to 7) with cluster-specific trajectories specified through the mixture parameter (mixture = ~ ns(CalendarAge, df)). Model fitting employed maximum likelihood estimation with convergence assessed using default lcmm criteria (log-likelihood change < 0.0001) and maximum 1000 iterations. Different residual variances per cluster were permitted (nwg = TRUE) to accommodate heteroscedasticity across latent classes, with base model parameters serving as starting values (B = base_model) for optimization stability.

For each model combination, comprehensive evaluation metrics were computed including information criteria (AIC and BIC) for model comparison with complexity penalties, and multiple clustering validation metrics including silhouette width, Dunn Index, Davies-Bouldin Index, and Calinski-Harabasz Index. Individual-level summary statistics were computed for distance calculations incorporating chronological age range, mean outcome measure trajectory values, and individual slope estimates per player, with all features normalized to ensure equal contribution to clustering validation metrics.

The same weighted consensus approach was applied as in the hybrid method, prioritizing metrics appropriate for trajectory modelling: BIC (30% weight), Silhouette coefficient (30% weight), and AIC (25% weight), with traditional distance-based separation metrics receiving moderate to low weighting: Calinski-Harabasz Index (10% weight), Dunn Index (2% weight), and Davies-Bouldin Index (3% weight).

#### Model Selection Process

Model selection balanced statistical fit with practical interpretability, with optimal cluster number and spline complexity chosen based on systematic evaluation of the consensus scoring approach across all model combinations.

#### Cluster Assignment Process

Individual cluster membership was determined using maximum posterior probabilities from the fitted model with hard cluster assignments based on the highest probability class for each individual player.

#### Model Diagnostics Process

Comprehensive model diagnostics were implemented through systematic diagnostic visualizations and summary reports paralleling the hybrid approach methodology. Cluster size distribution was evaluated through bar plots showing the number and percentage of observations per cluster.

Posterior probability analysis included extraction of individual posterior probabilities for soft clustering assignments, calculation of maximum posterior probability and uncertainty measures (1 - max posterior), with box plots displaying uncertainty distributions by cluster using established certainty thresholds (uncertainty ≤0.2 indicating high certainty assignments).

Distance-based validation employed Euclidean distance matrices from scaled individual-level features, with comprehensive silhouette analysis including cluster-specific and overall average silhouette widths displayed through bar plots with quality thresholds: ≥0.7 excellent, ≥0.5 good, ≥0.25 moderate, <0.25 poor.

Separation metrics were systematically computed and displayed including the Dunn Index with quality thresholds (≥1.0 excellent, ≥0.5 good, <0.5 poor separation), Davies-Bouldin Index with compactness benchmarks ($\leq 1.0$ excellent, $\leq 2.0$ good, $\geq 2.0$ poor), and Calinski-Harabasz Index for between-cluster to within-cluster variance ratio assessment.

Bootstrap stability assessment was conducted through 30 resampling iterations with 80% data subsampling, with models refitted and stability quantified using Adjusted Rand Index between original and bootstrap cluster assignments. Histogram distributions showed stability results with established thresholds: $\geq 0.8$ excellent, $\geq 0.6$ good, $\geq 0.4$ moderate, $<0.4$ poor stability.

Residual diagnostics were implemented through scatter plots showing residuals vs fitted values colored by cluster for homoscedasticity assessment, and faceted histograms displaying residual frequency distributions per cluster for normality evaluation and cluster-specific residual pattern identification.

Bootstrap-based confidence intervals were created through 30 resampling iterations for trajectory visualization, providing model uncertainty quantification and enhancing interpretation of developmental pattern estimates.

Entropy-based separation analysis was calculated using Shannon entropy formula with quality assessment thresholds: $\leq 0.2$ excellent separation (clear cluster boundaries), 0.2-0.4 good separation, 0.4-0.6 moderate separation (some ambiguous cases), $>0.6$ poor separation (significant cluster overlap).

Comprehensive summary reports provided multi-dimensional quality assessment combining all diagnostic measures with overall model quality ratings and targeted improvement recommendations based on metric consensus across the validation framework.

### Supplementary 2. Results from the Latent Class Mixed Models (LCMM) clustering approach

Latent Class Mixed Models (LCMM) were fitted as a comparative methodological approach to the two-step hybrid framework (GAM + Mclust). Based on systematic model selection using weighted consensus scoring across multiple validation criteria (see Supplementary 1), the optimal LCMM solution identified df = 5 with ng = 6 clusters for vIANT, and df = 5 with ng = 6 clusters for v4. Comprehensive model diagnostics and quality assessment results for the LCMM approach are provided in Supplementary 3.

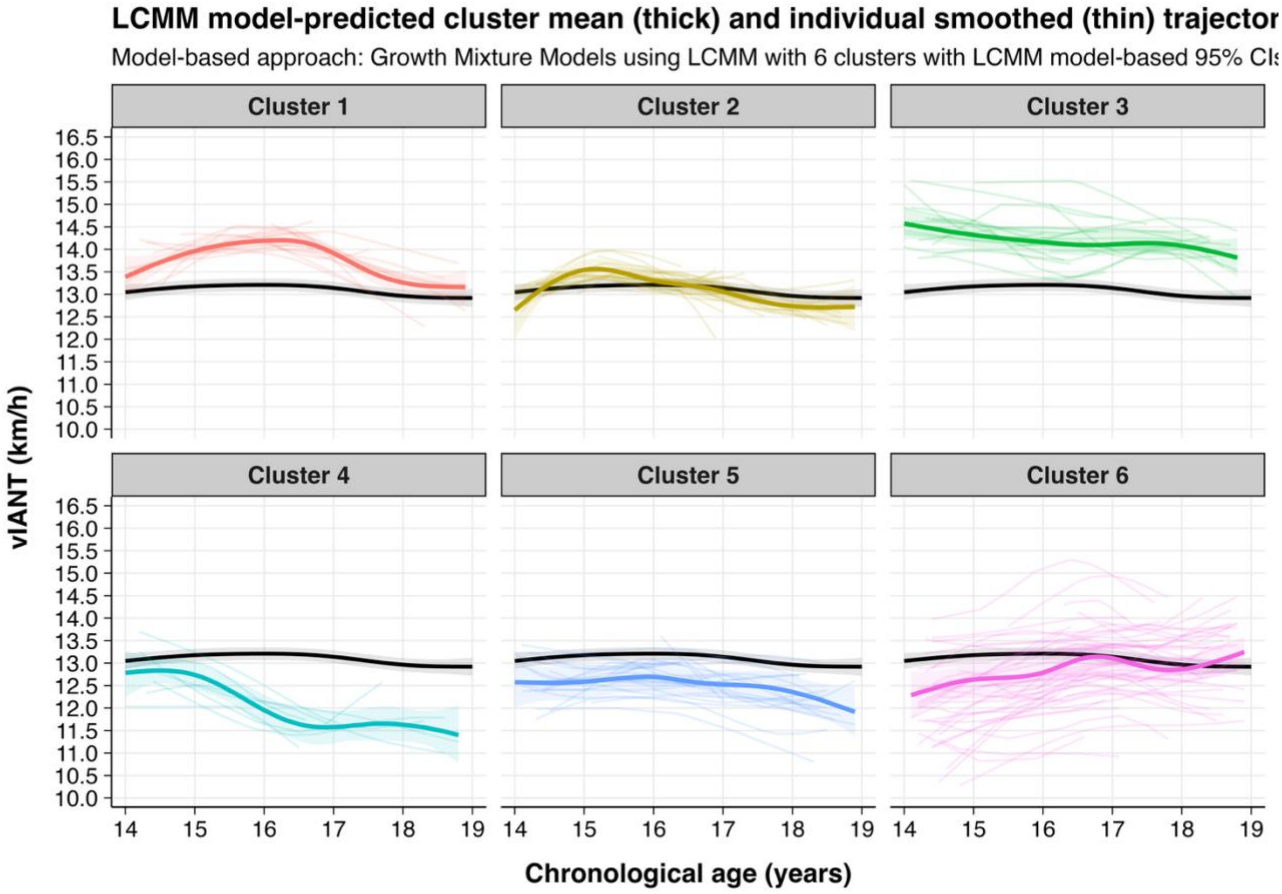

**Supplementary 2. Figure 1.** Distinct trajectory clusters for the outcome measure *velocity at the individual anaerobic threshold (vIANT)* derived from the Latent Class Mixed Model (LCMM) clustering approach. Each cluster shows individual trajectories and GAM model-predicted cluster means with model-based 95% confidence intervals. The black line represents the mean trajectory of the entire sample as reference with 95% confidence intervals.

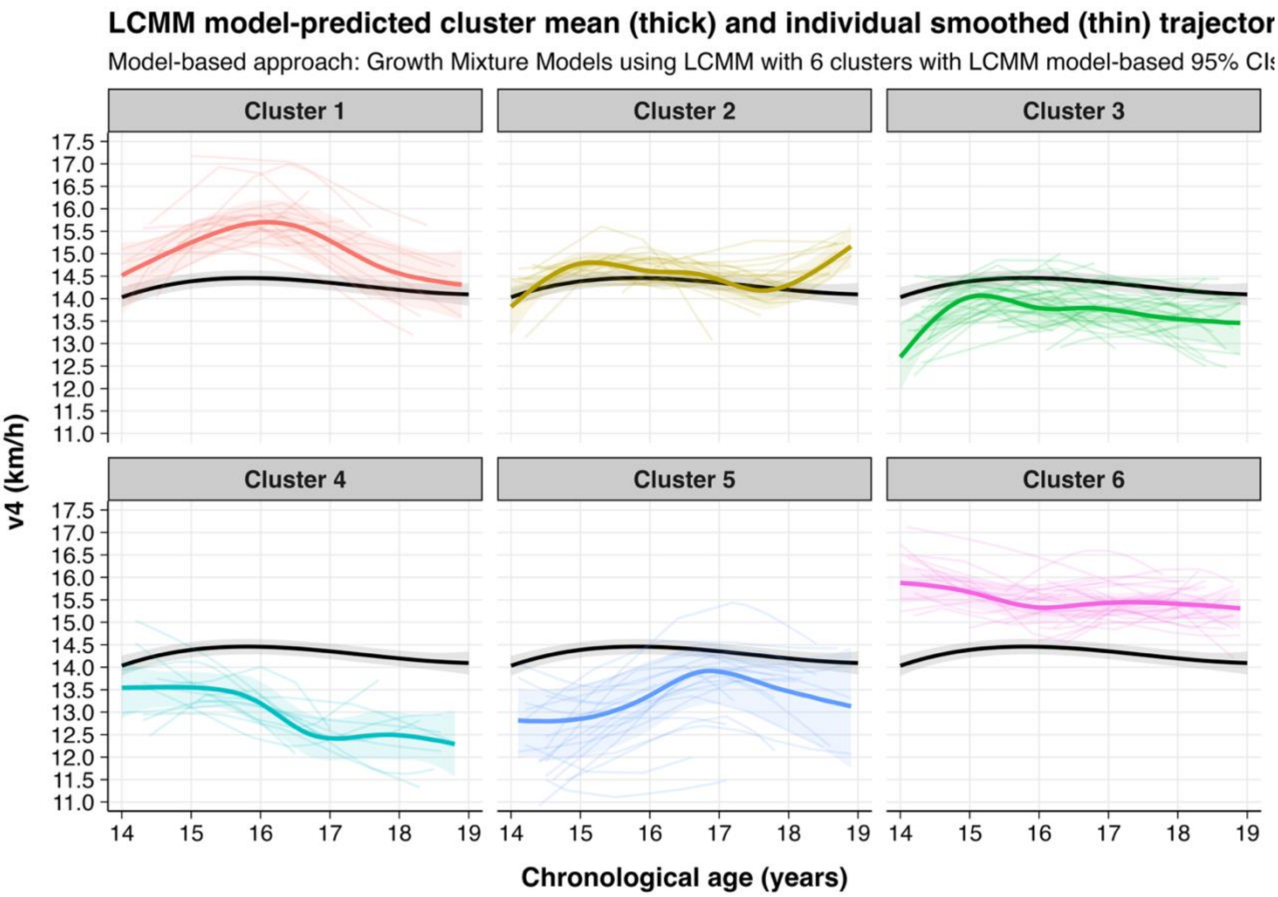

**Supplementary 2. Figure 2.** Distinct trajectory clusters for the outcome measure *velocity at lactate concentration of 4 mmol.L⁻¹ (v4)* derived from the Latent Class Mixed Model (LCMM) clustering approach, showing individual trajectories and GAM model-predicted cluster means with model-based 95% confidence intervals. The black line represents the mean trajectory of the entire sample as reference with 95% confidence intervals.

**Supplementary 3. Comprehensive model diagnostics for vIANT of both clustering approaches (Hybrid approach (GAM + Mclust) and Latent Class Mixed Model (LCMM))**

***Model diagnostics for hybrid approach (GAM + Mclust)***

<u>Step 1: Individual trajectory estimation (GAM model diagnostics)</u>

Supplementary 3. Table 1. Model selection results for the Generalized Additive Model (GAM) across k-values.

| k-value | Deviance explained | R squared | AIC | BIC | REML score | edf | converged |
|---|---|---|---|---|---|---|---|
| 3 | 0.824 | 0.783 | 1970 | 3378 | 1214 | 265 | TRUE |
| 4 | 0.829 | 0.786 | 1964 | 3455 | 1218 | 280 | TRUE |
| 5 | 0.832 | 0.787 | 1964 | 3514 | 1228 | 292 | TRUE |
| 6 | 0.838 | 0.791 | 1953 | 3608 | 1242 | 312 | TRUE |
| 7 | 0.836 | 0.791 | 1946 | 3542 | 1222 | 301 | TRUE |

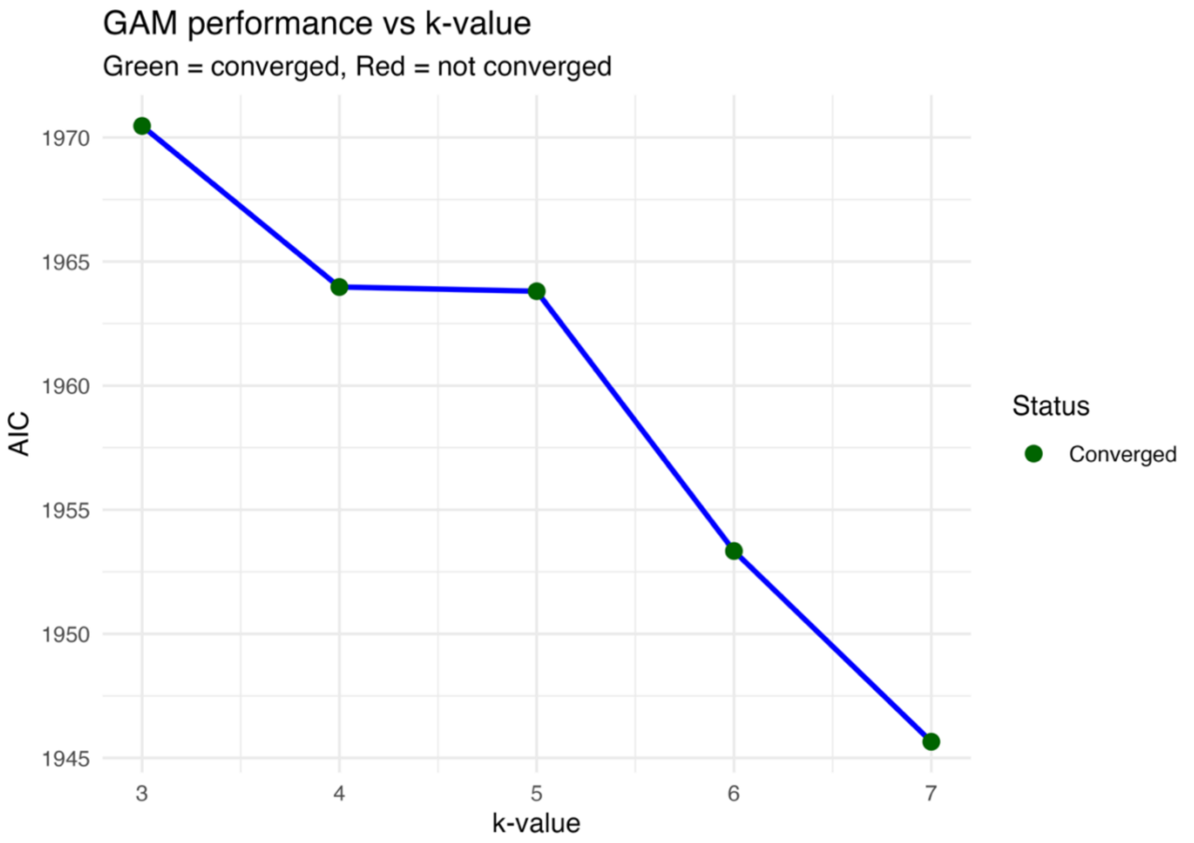

Supplementary 3. Figure 1. AIC values derived from Generalized Additive Models (GAM) across k-values 3 to 7.

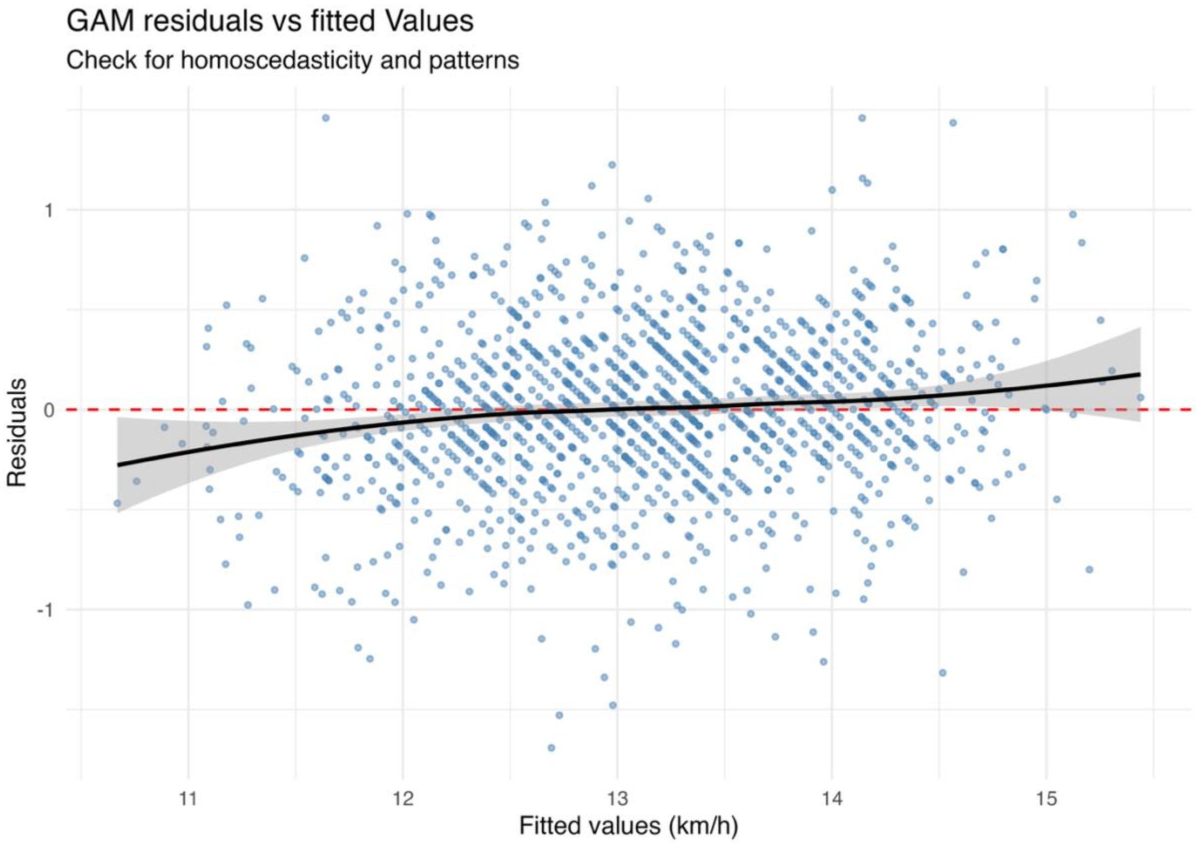

Supplementary 3. Figure 2. Residuals versus fitted values from the Generalized Additive Model (GAM).

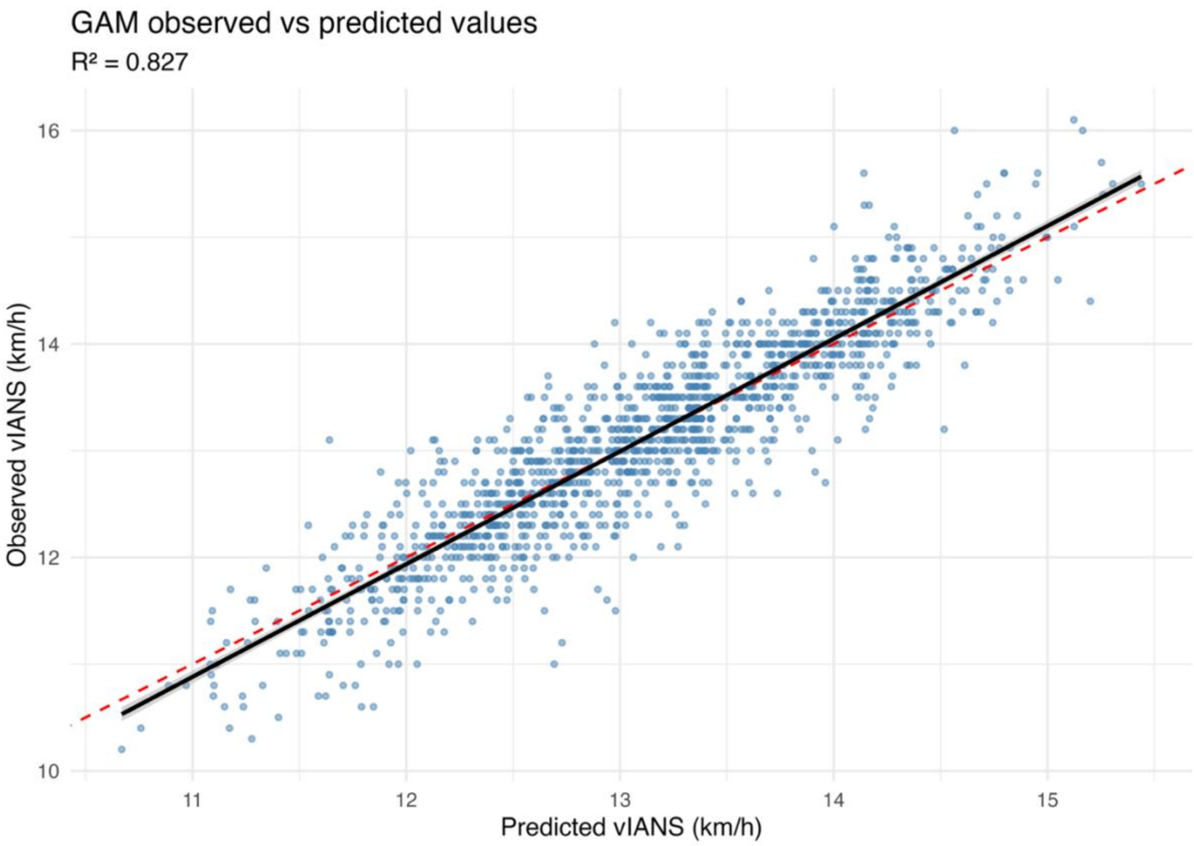

Supplementary 3. Figure 3. Observed versus predicted values from the Generalized Additive Model (GAM).

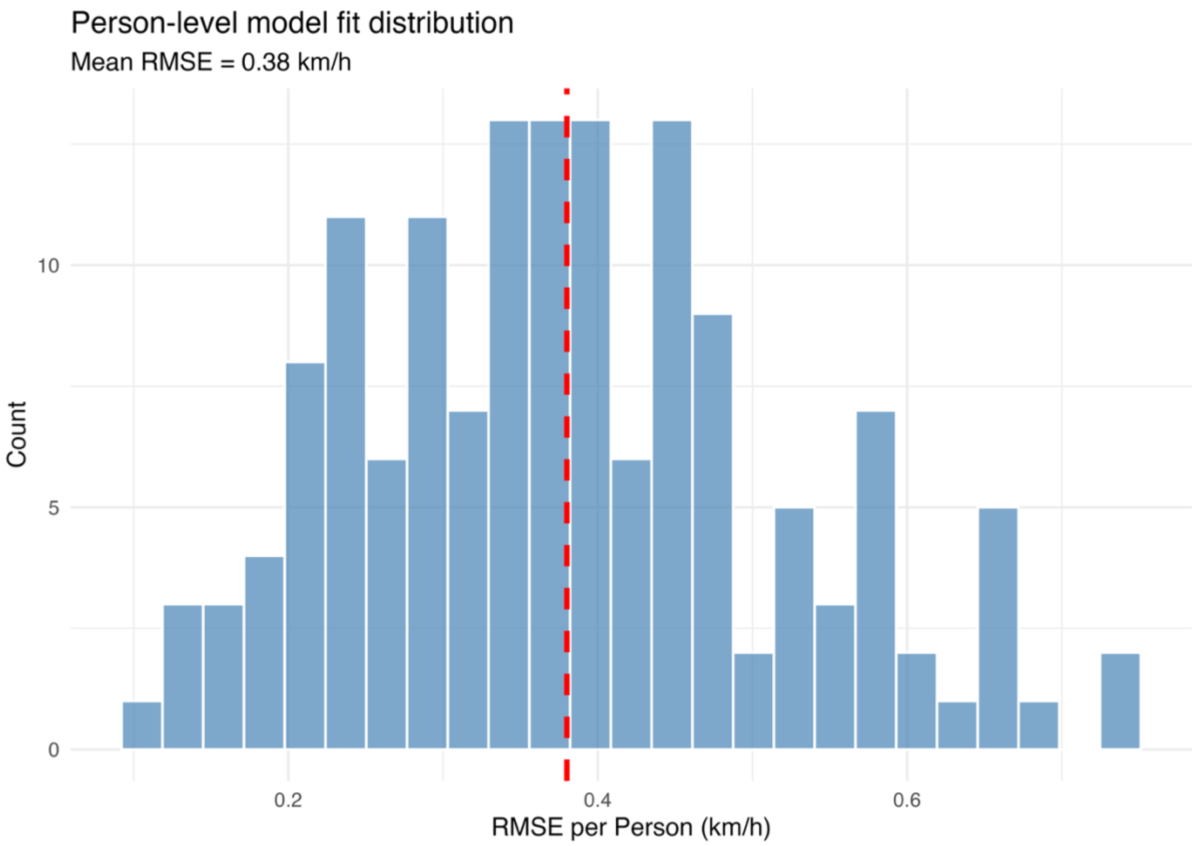

Supplementary 3. Figure 4. Distribution of player-level fit indices in the Generalized Additive Model (GAM).

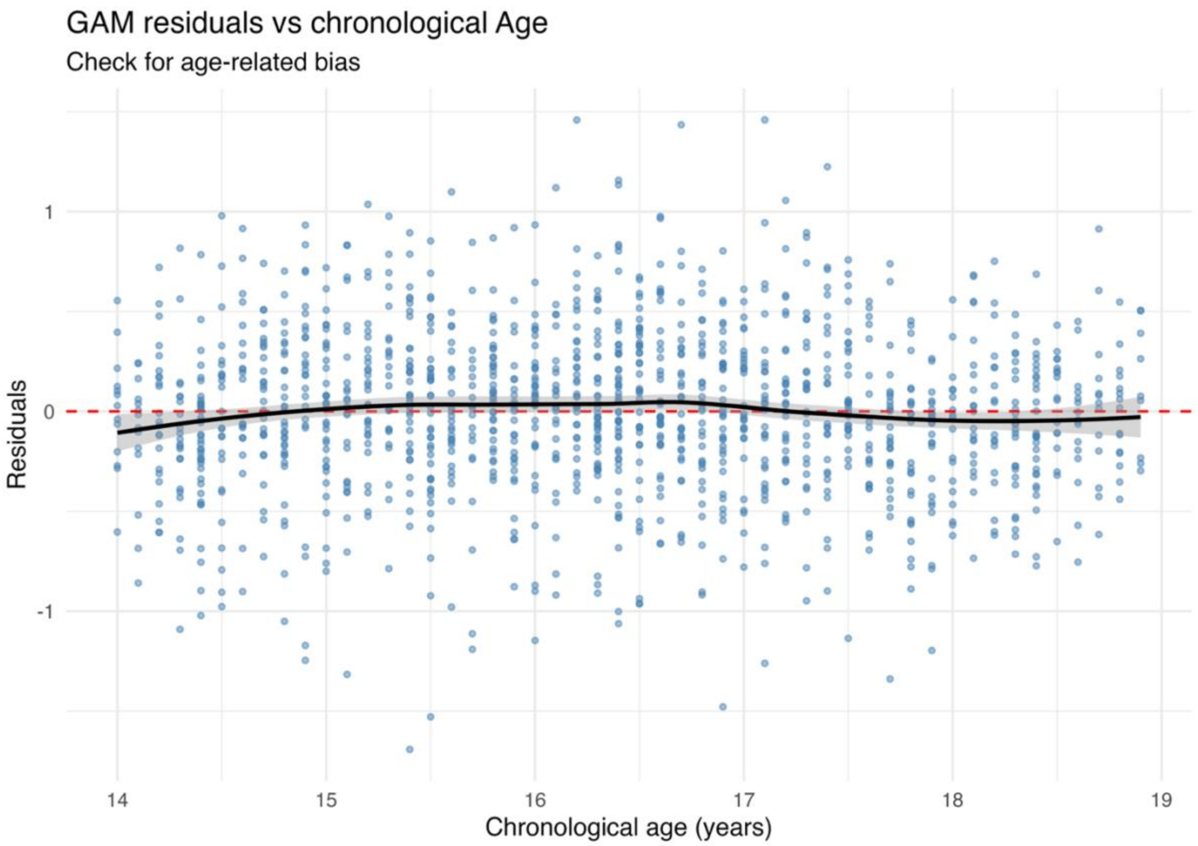

Supplementary 3. Figure 5. Residuals versus chronological age for the Generalized Additive Model (GAM).

Step 2: Feature-Based clustering (Mclust diagnostics)

Supplementary 3. Table 2. Mclust model selection results across cluster numbers and covariance structures.

| Number clusters | Model | BIC | AIC | loglik | Dunn | DB | CH | Silhouette |
|---|---|---|---|---|---|---|---|---|
| 6 | VEI | -4838 | 4492 | -2131 | 0.081 | 1.4 | 30.2 | 0.129 |
| 5 | VEI | -4959 | 4665 | -2234 | 0.097 | 1.3 | 33.2 | 0.159 |
| 4 | VEI | -5073 | 4830 | -2334 | 0.031 | 1.5 | 41.0 | 0.207 |
| 3 | EVI | -5370 | 5100 | -2460 | 0.066 | 1.4 | 46.8 | 0.234 |
| 3 | VVI | -5476 | 5200 | -2508 | 0.068 | 1.6 | 28.1 | 0.143 |
| 3 | VEI | -5531 | 5339 | -2605 | 0.024 | 1.7 | 24.5 | 0.121 |

Note: G refers to the number of clusters estimated in the model

Supplementary 3. Table 3: Final Mclust model quality metrics and interpretations.

| Metric | Value | Interpretation |
|---|---|---|
| Average Silhouette Width | 0.207 | Poor |
| Dunn | 0.880 | Good |
| Davies-Bouldin (DB) | 2.1 | Poor |
| Calinski-Harabasz (CH) | 15.1 | Higher is better |
| Bootstrap Stability (Mean ARI) | 0.59 | Moderate |
| Bootstrap Stability (SD) | 0.15 | - |
| Entropy | 0.02 | Good |
| Mean Uncertainty | 0.01 | Good |

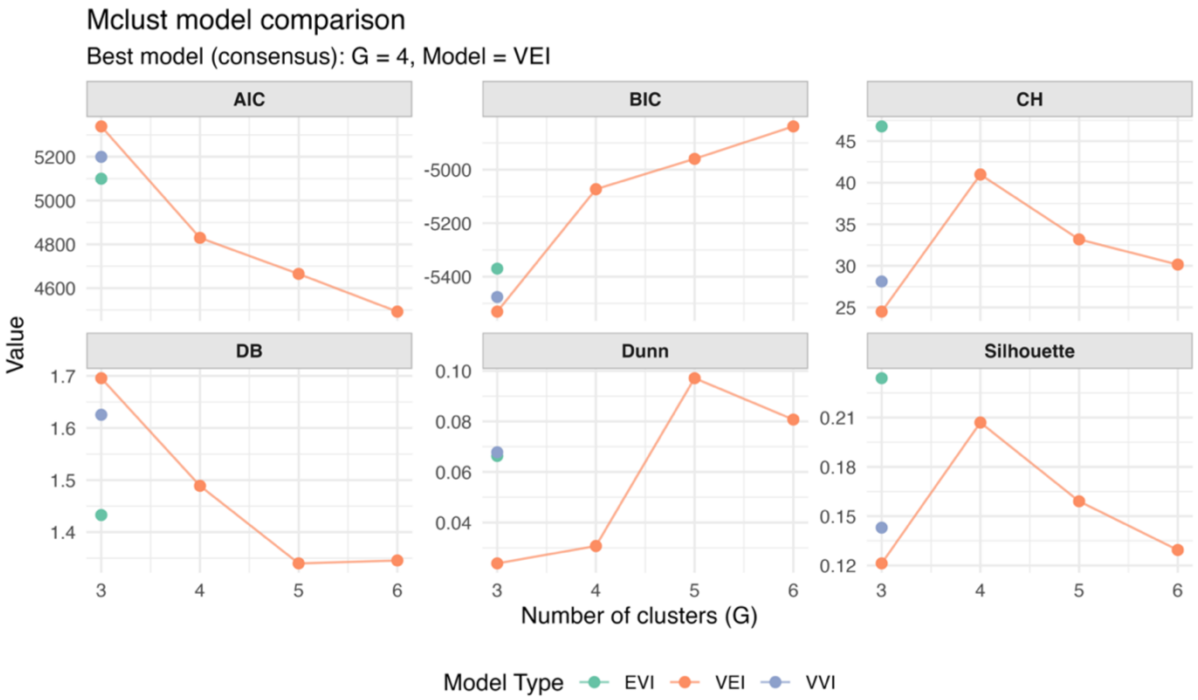

Supplementary 3. Figure 6. Comparison of Mclust model selection metrics across cluster numbers and covariance structures. CH: Calinski-Harabasz; DB: Davies-Bouldin.

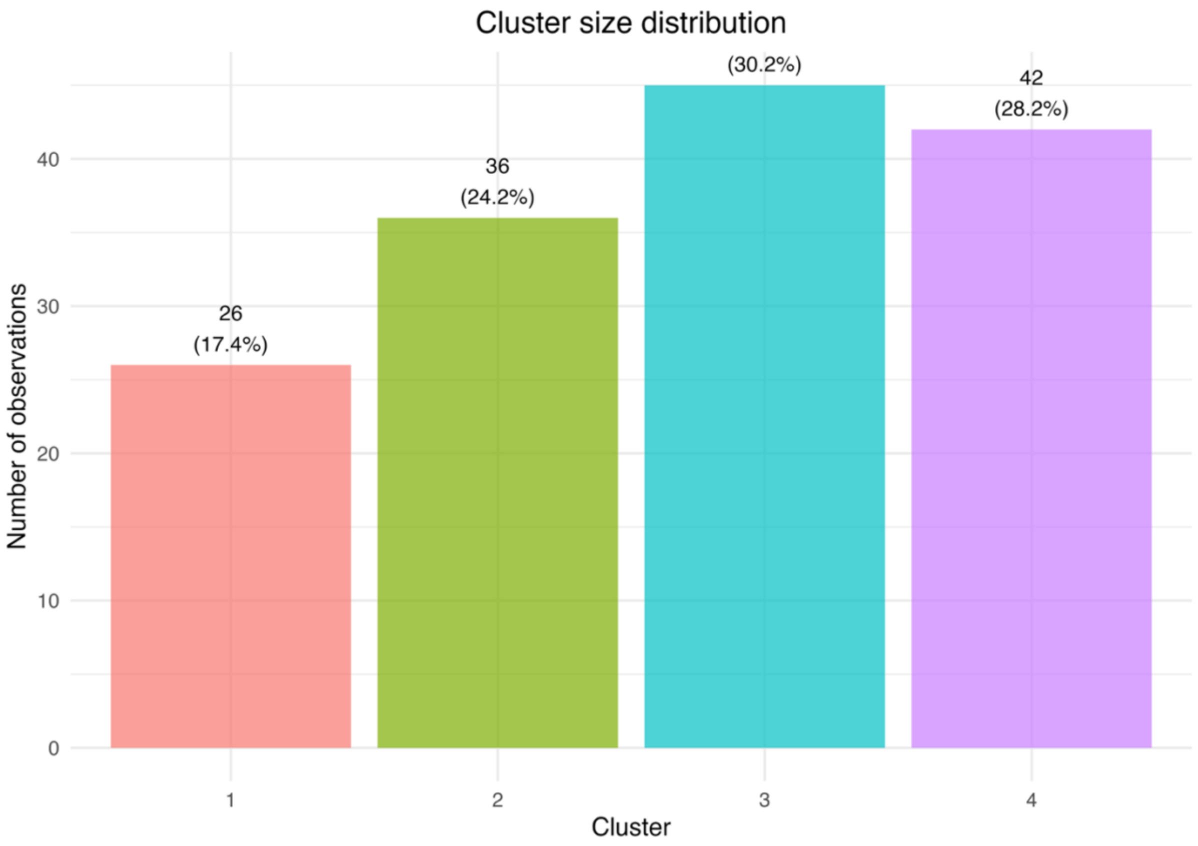

Supplementary 3. Figure 7. Cluster size distribution of the selected Mclust model.

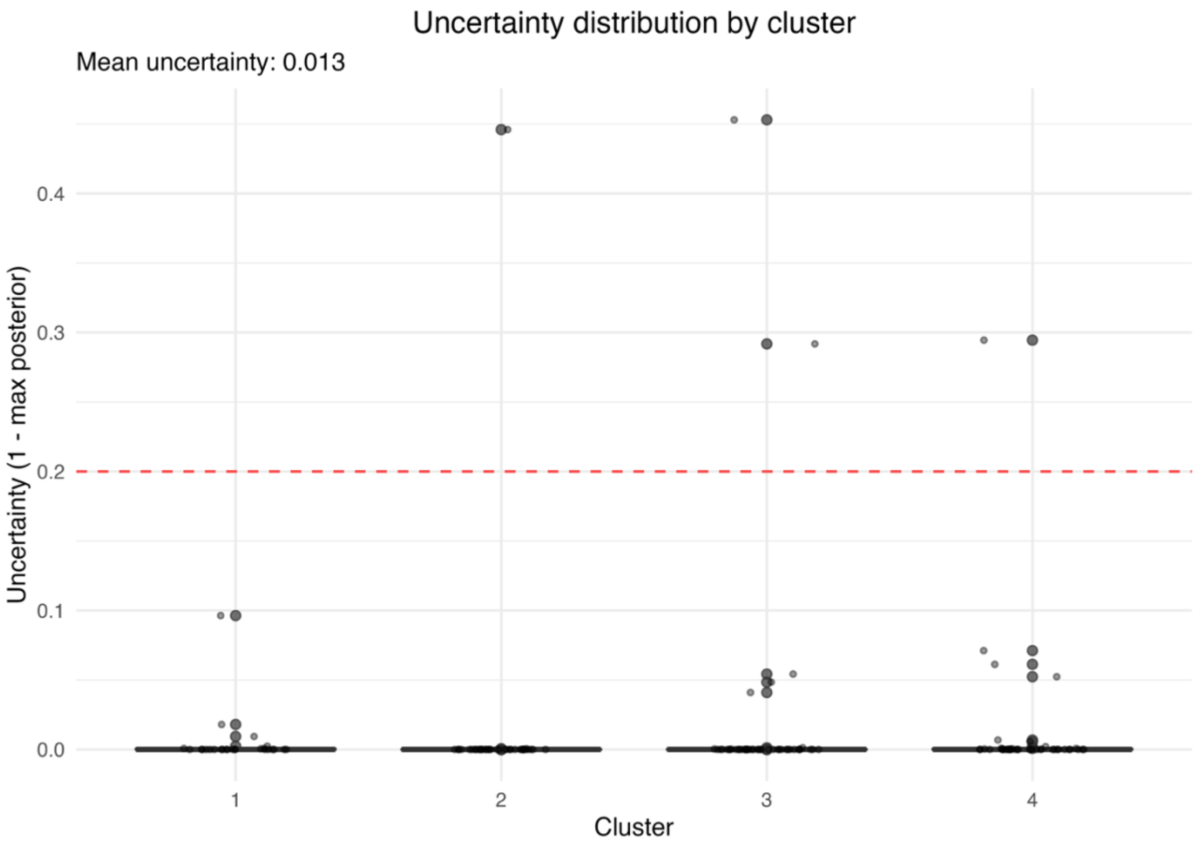

Supplementary 3. Figure 8. Distribution of classification uncertainty by cluster of the selected Mclust model.

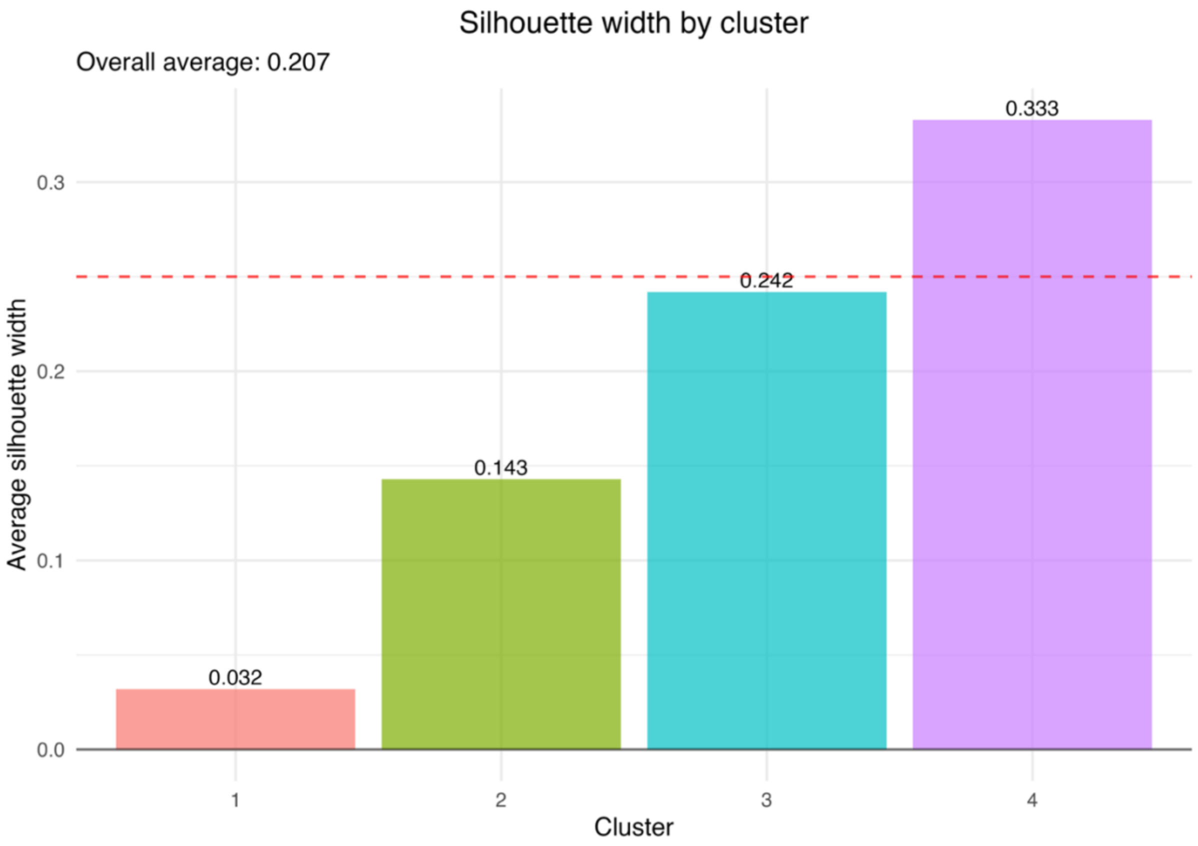

Supplementary 3. Figure 9. Silhouette width by cluster of the selected Mclust model.

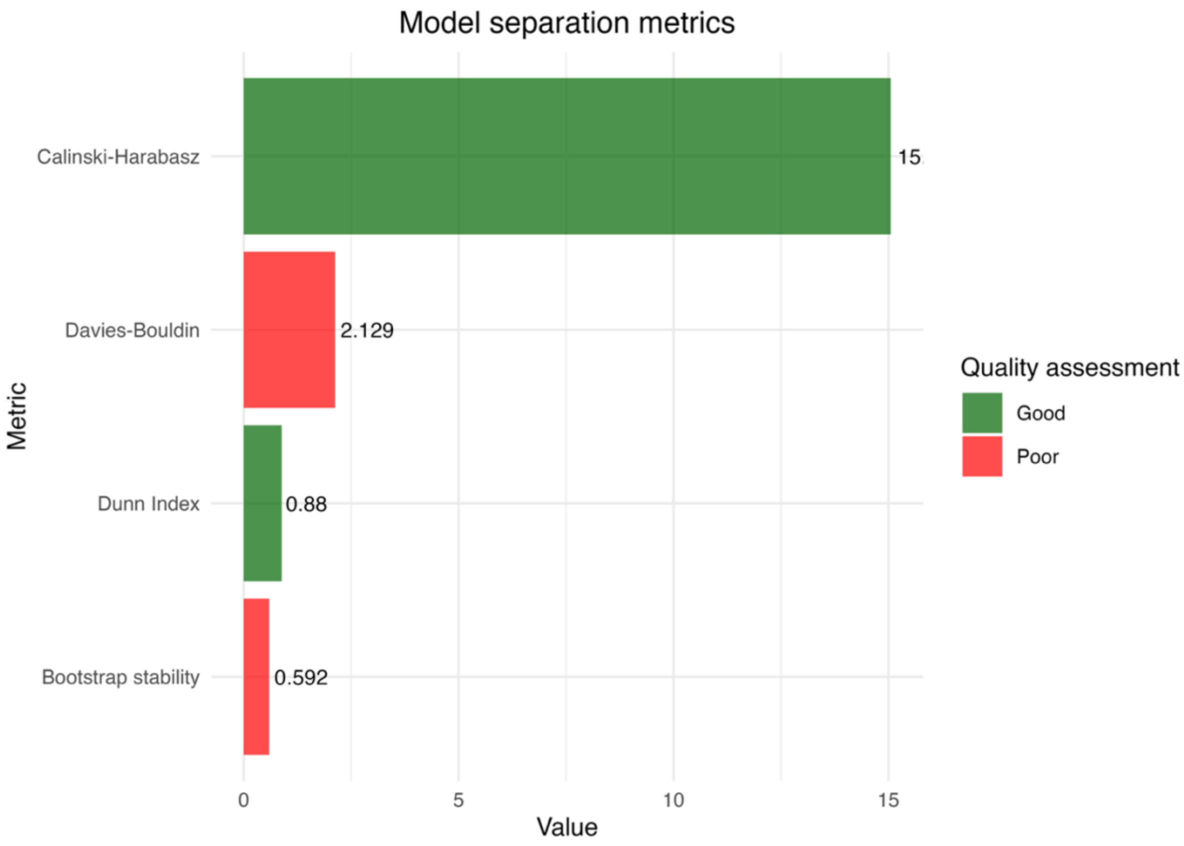

Supplementary 3. Figure 10. Comparison of separation metrics (Calinski-Harabasz, Davies-Bouldin, Dunn and Bootstrap stability) of the selected Mclust model.

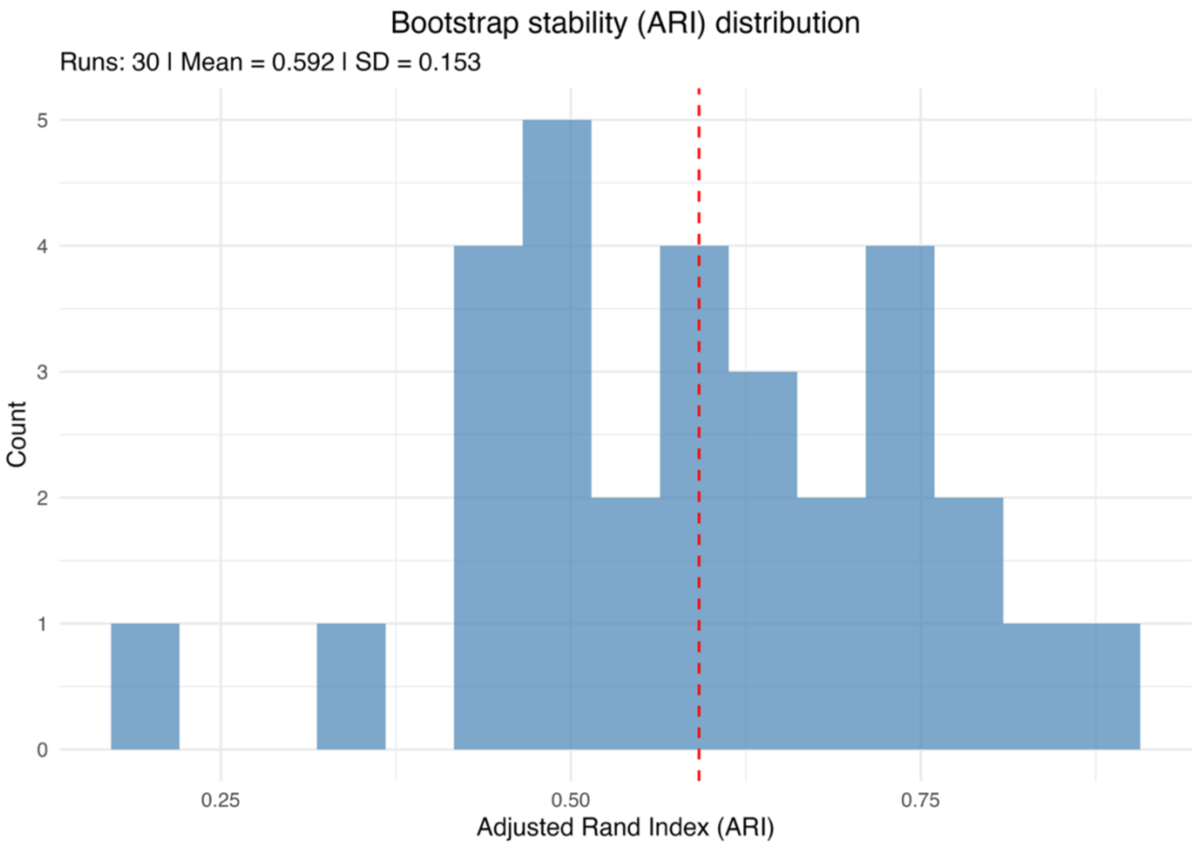

Supplementary 3. Figure 11. Bootstrap distribution of Adjusted Rand Index (ARI) values assessing cluster stability of the selected Mclust model.

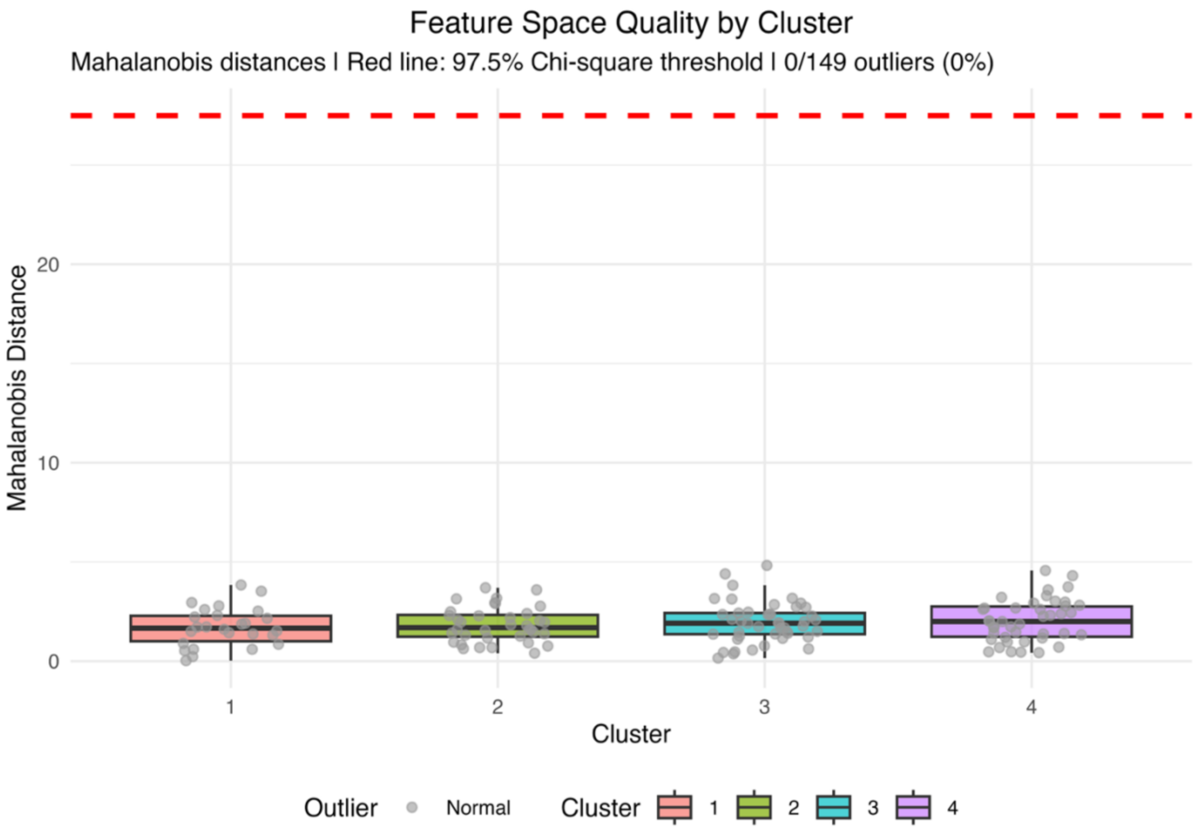

Supplementary 3. Figure 12. Mahalanobis distance by cluster representing feature space separation of the selected Mclust model.

#### ***Model diagnostics for Latent Class Mixed Model (LCMM) approach***

<u>Latent Class Mixed Model (LCMM) selection diagnostics</u>

Supplementary 3. Table 4. Comprehensive Latent Class Mixed Model (LCMM) comparison across degrees of freedom (df) and cluster numbers (ng) for all computed models.

| Model | df | ng | loglik | AIC | BIC | Dunn | Davies-Bouldin (DB) | Calinski Harabasz (CH) | Silhouette | converged |
|---|---|---|---|---|---|---|---|---|---|---|
| df3_ng3 | 3 | 3 | -1190 | 2427 | 2496 | 0.003 | 4.0 | 4.0 | -0.104 | TRUE |
| df4_ng3 | 4 | 3 | -1188 | 2427 | 2505 | 0.003 | 3.6 | 5.1 | -0.086 | TRUE |
| df5_ng3 | 5 | 3 | -1184 | 2426 | 2513 | 0.003 | 3.8 | 4.3 | -0.108 | TRUE |
| df3_ng4 | 3 | 4 | -1193 | 2444 | 2531 | 0.003 | 6.5 | 3.0 | -0.219 | TRUE |
| df4_ng4 | 4 | 4 | -1189 | 2445 | 2544 | 0.003 | 29.0 | 2.9 | -0.224 | TRUE |
| df4_ng5 | 4 | 5 | -1172 | 2424 | 2544 | 0.003 | 9.4 | 5.4 | -0.207 | TRUE |
| df3_ng6 | 3 | 6 | -1171 | 2425 | 2548 | 0.003 | 9.3 | 5.0 | -0.204 | TRUE |
| df5_ng5 | 5 | 5 | -1162 | 2415 | 2550 | 0.003 | 25.3 | 2.6 | -0.315 | TRUE |

| | | | | | | | | | | |
|---|---|---|---|---|---|---|---|---|---|---|
| df3_ng5 | 3 | 5 | -1189 | 2449 | 2554 | 0.003 | 5.4 | 2.4 | -0.248 | TRUE |
| df5_ng4 | 5 | 4 | -1186 | 2447 | 2558 | 0.003 | 33.5 | 3.1 | -0.219 | TRUE |
| df4_ng6 | 4 | 6 | -1168 | 2431 | 2572 | 0.004 | 2.9 | 15.3 | -0.145 | FALSE |
| df5_ng6 | 5 | 6 | -1158 | 2422 | 2581 | 0.004 | 4.1 | 12.3 | -0.101 | TRUE |

Note: df refers to the degrees of freedom of the natural spline used to model the trajectory shape, and ng refers to the number of latent classes (clusters) estimated in the model.

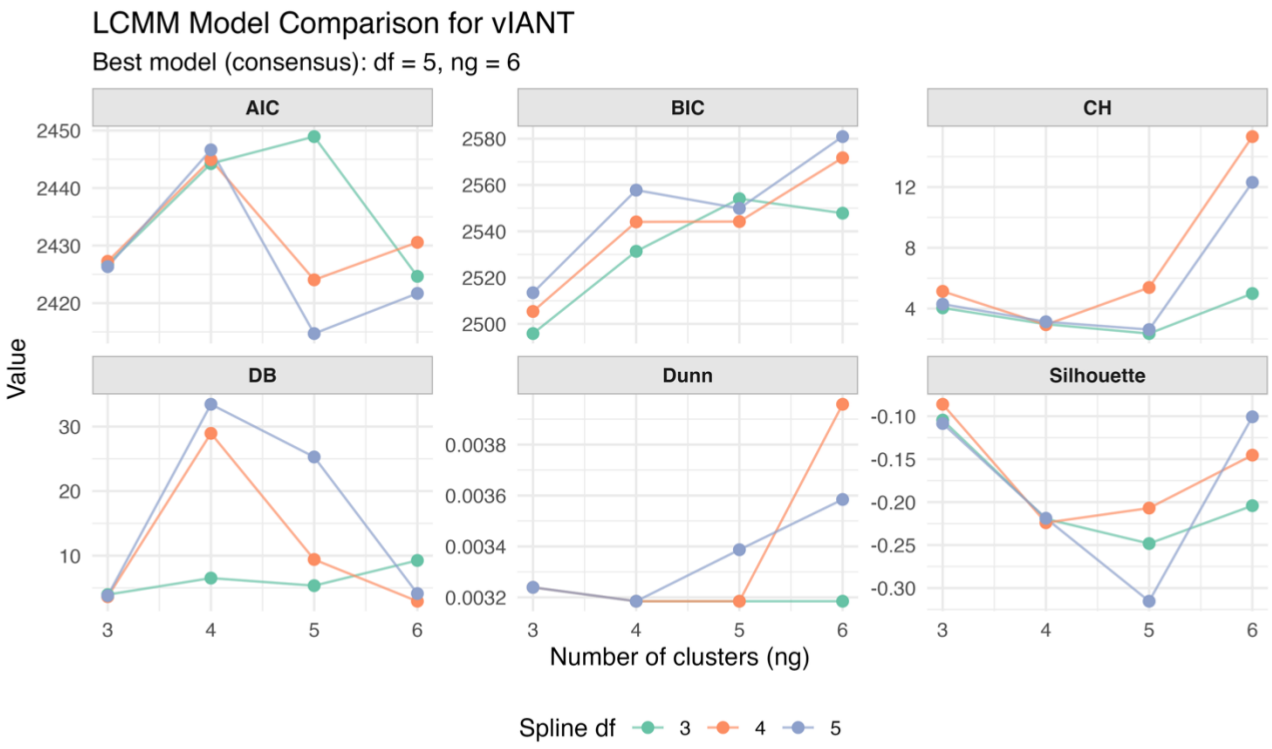

Supplementary 3. Figure 13. Comparison of Latent Class Mixed Model (LCMM) selection metrics across degrees of freedom (df) and cluster numbers (ng). CH: Calinski-Harabasz; DB: Davies-Bouldin

Latent Class Mixed Model (LCMM) clustering quality diagnostics

Supplementary 3. Table 5. Final Latent Class Mixed Model (LCMM) clustering quality metrics and interpretations.

| Metric | Value | Interpretation |
|---|---|---|
| Average Silhouette Width | -0.002 | Poor |
| Dunn | 0.831 | Good |
| Davies-Bouldin (DB) | 4.9 | Poor |

| | | |
|---|---|---|
| Calinski-Harabasz (CH) | 7.8 | Higher is better |
| Bootstrap Stability (Mean ARI) | 0.22 | Moderate |
| Bootstrap Stability (SD) | 0.11 | - |
| Entropy | 0.26 | Good |
| Mean Uncertainty | 0.19 | Good |

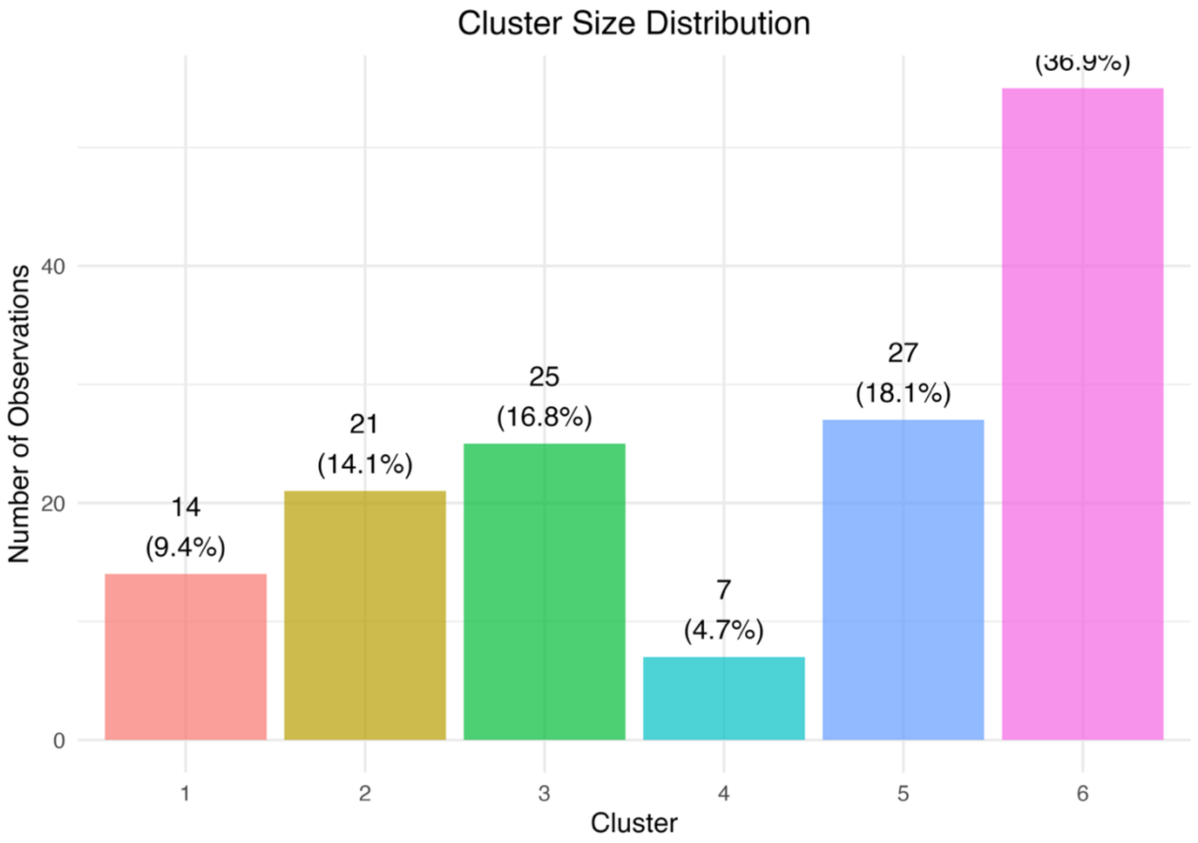

Supplementary 3. Figure 14. Cluster size distribution of the selected Latent Class Mixed Model (LCMM).

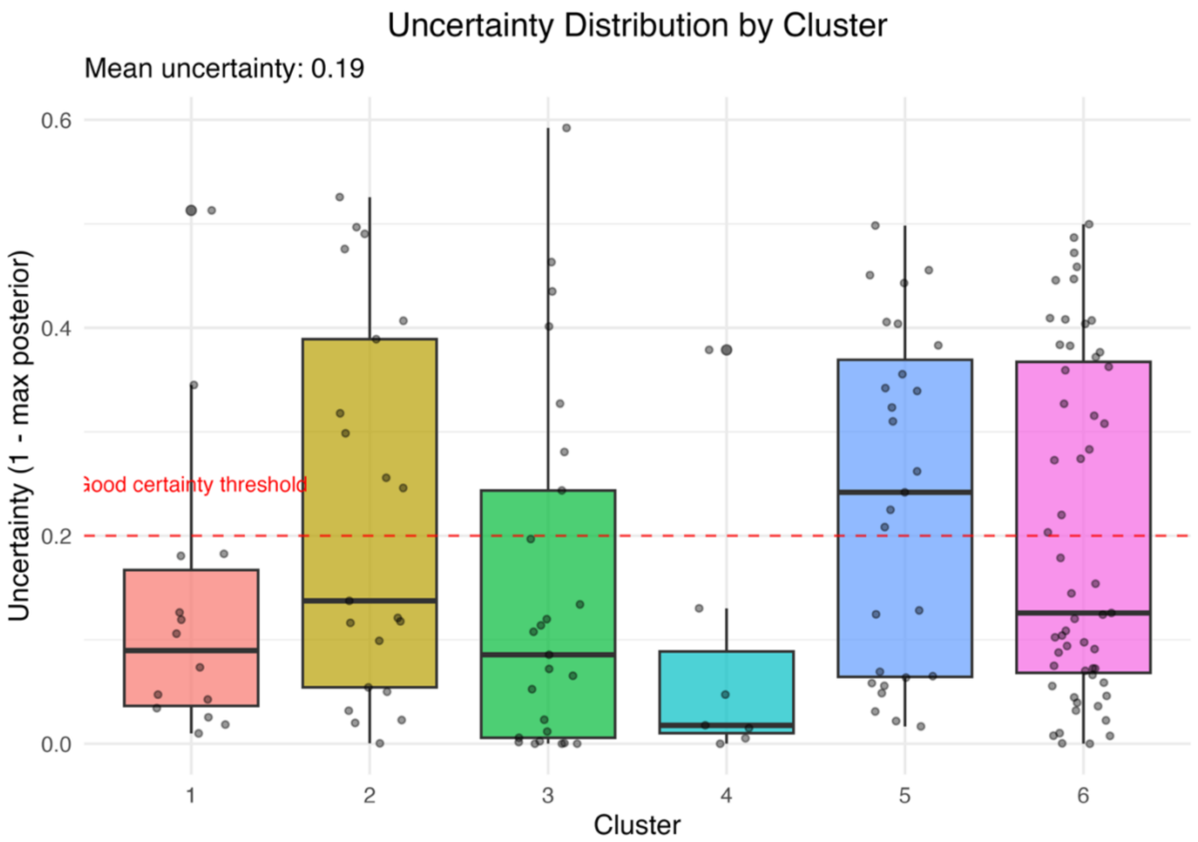

Supplementary 3. Figure 15. Distribution of classification uncertainty by cluster of the selected Latent Class Mixed Model (LCMM).

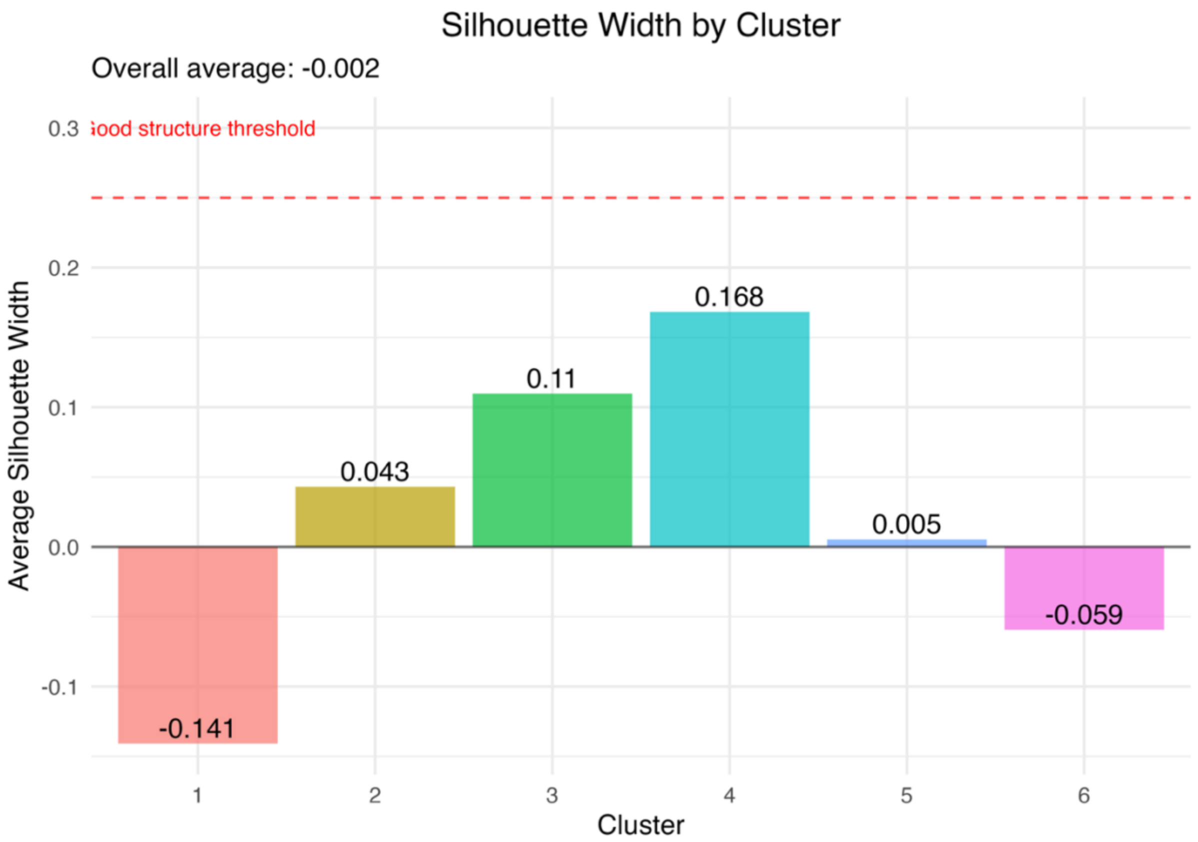

Supplementary 3. Figure 16. Silhouette width by cluster of the selected Latent Class Mixed Model (LCMM).

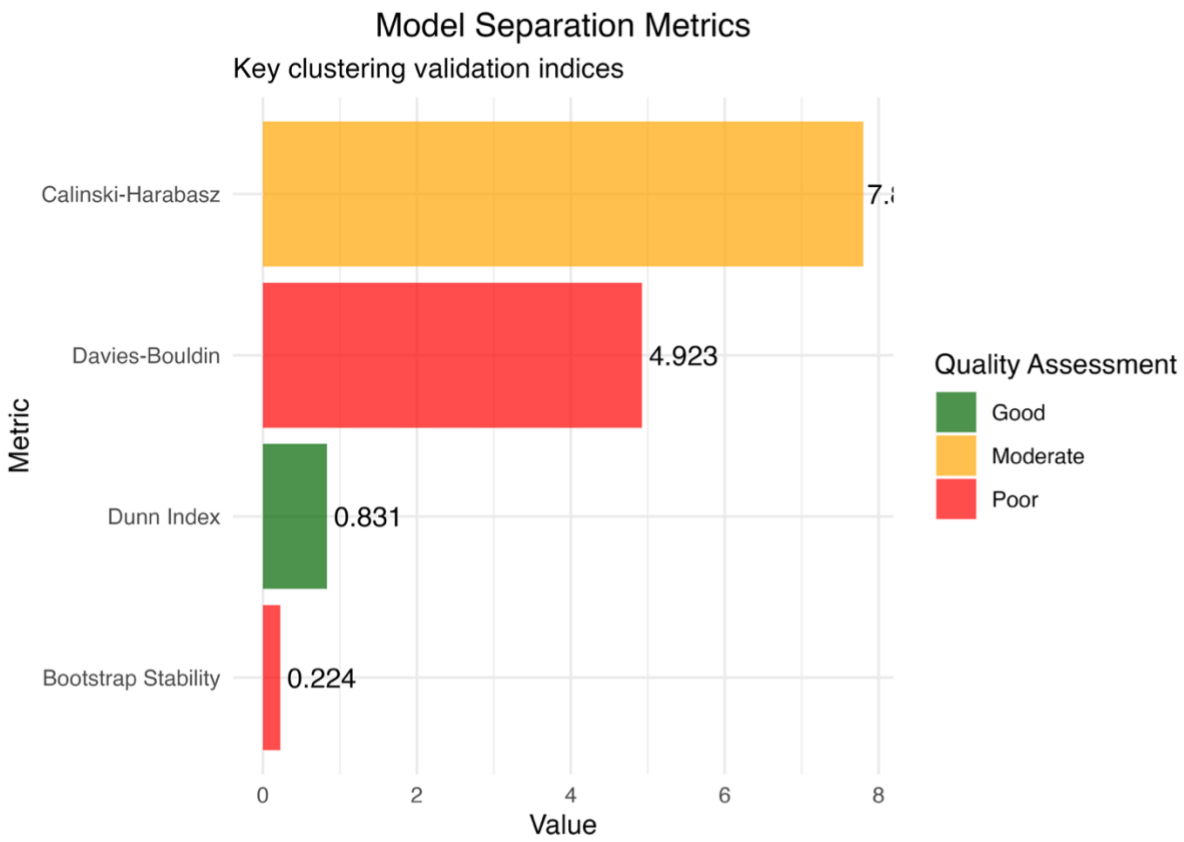

Supplementary 3. Figure 17. Comparison of separation metrics (Calinski-Harabasz, Davies-Bouldin, Dunn and Bootstrap stability) of the selected Latent Class Mixed Model (LCMM).

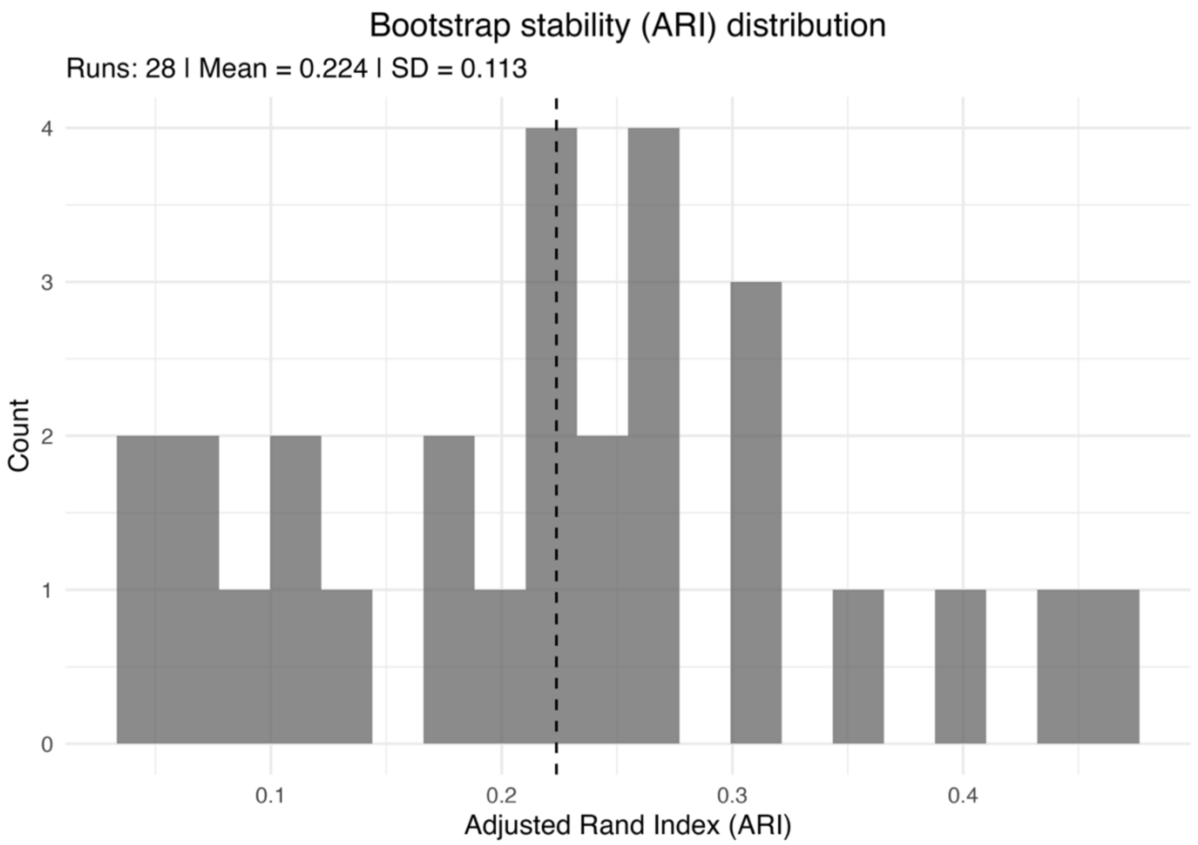

Supplementary 3. Figure 18. Bootstrap distribution of Adjusted Rand Index (ARI) values assessing cluster stability of the selected Latent Class Mixed Model (LCMM).

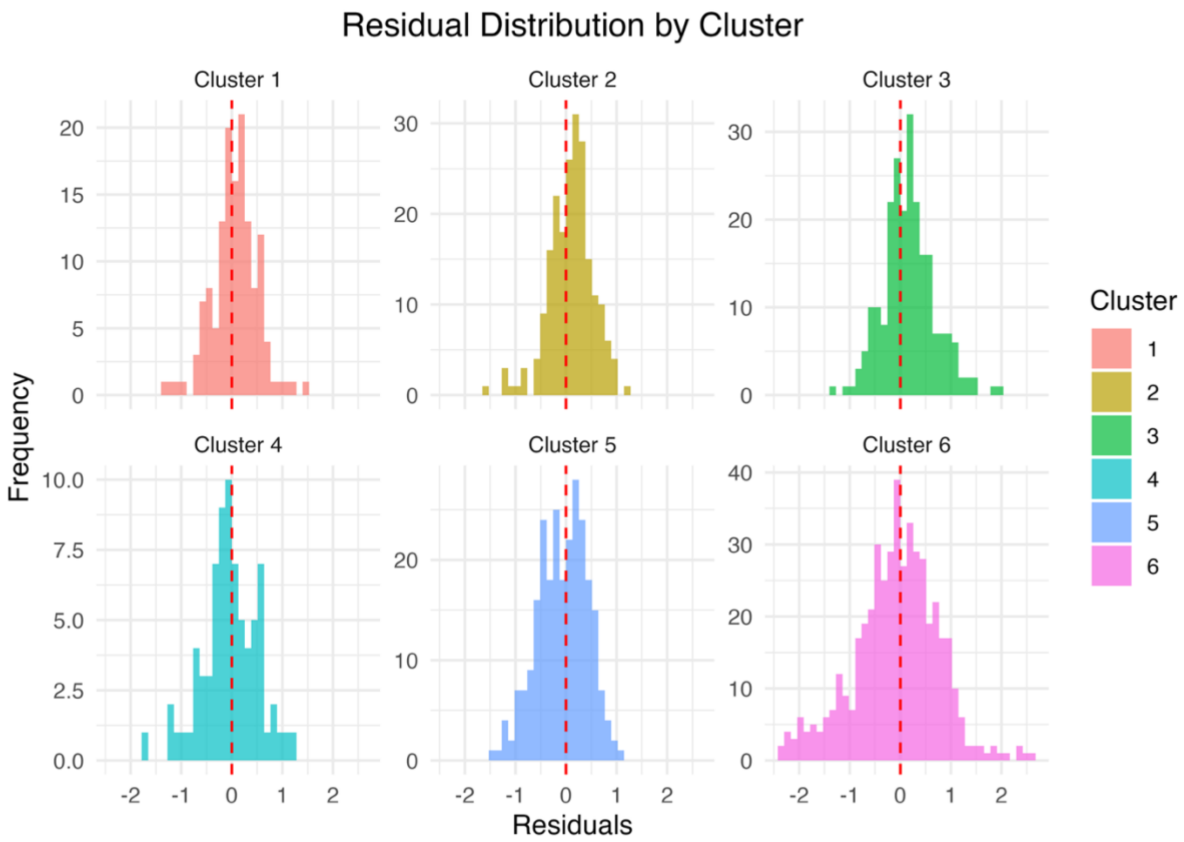

Supplementary 3. Figure 19. Distribution of residuals by cluster from the selected Latent Class Mixed Model (LCMM).

**Supplementary 4. Comprehensive model diagnostics for v4 of both clustering approaches (Hybrid approach (GAM + Mclust) and Latent Class Mixed Model (LCMM))**

***Model diagnostics for hybrid approach (GAM + Mclust)***

Step 1: Individual trajectory estimation (GAM model diagnostics)

Supplementary 4. Table 1. Model selection results for the Generalized Additive Model (GAM) across k-values.

| k-value | Deviance explained | R squared | AIC | BIC | REML score | edf | converged |
|---|---|---|---|---|---|---|---|
| 3 | 0.780 | 0.731 | 2876 | 4211 | 1637 | 251 | TRUE |
| 4 | 0.787 | 0.736 | 2866 | 4295 | 1639 | 268 | TRUE |
| 5 | 0.789 | 0.737 | 2871 | 4351 | 1648 | 278 | TRUE |
| 6 | 0.796 | 0.74 | 2868 | 4461 | 1665 | 300 | TRUE |
| 7 | 0.794 | 0.74 | 2858 | 4385 | 1646 | 287 | TRUE |

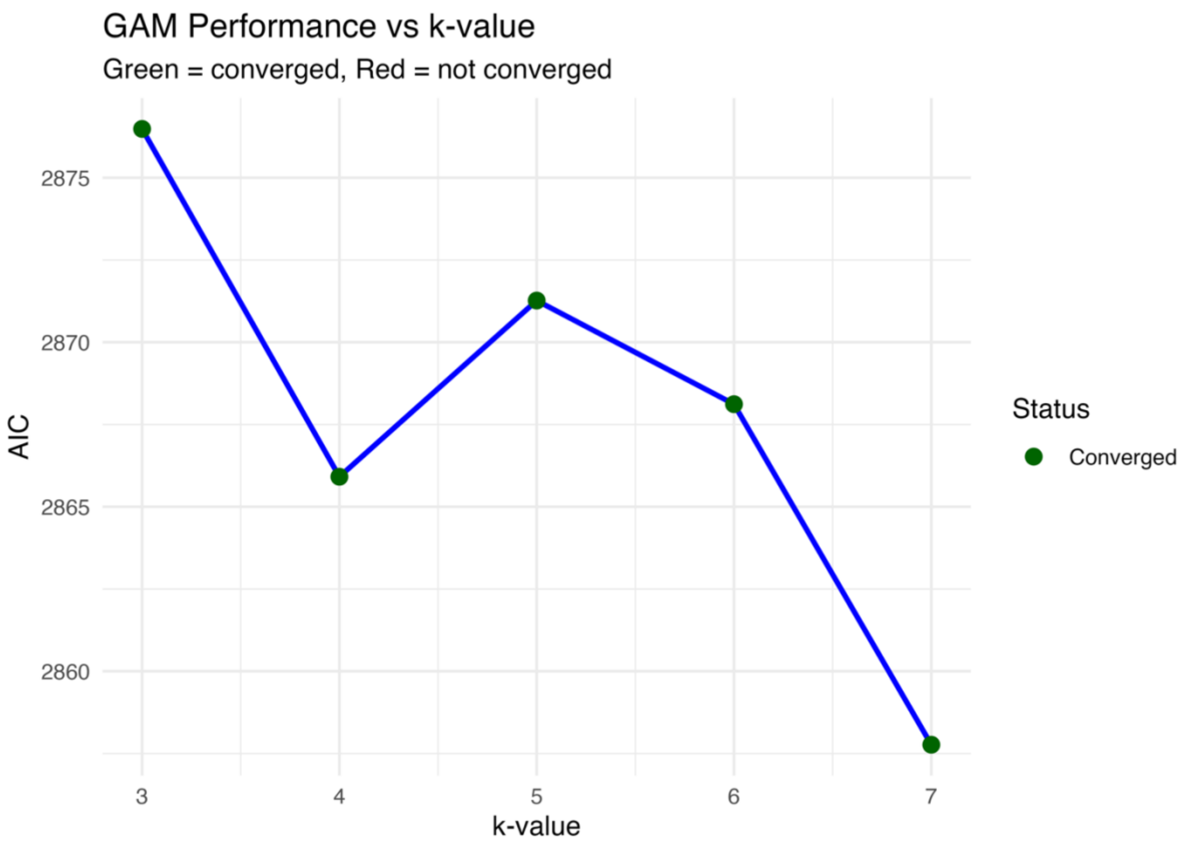

Supplementary 4. Figure 1. AIC values derived from Generalized Additive Models (GAM) across k-values 3 to 7.

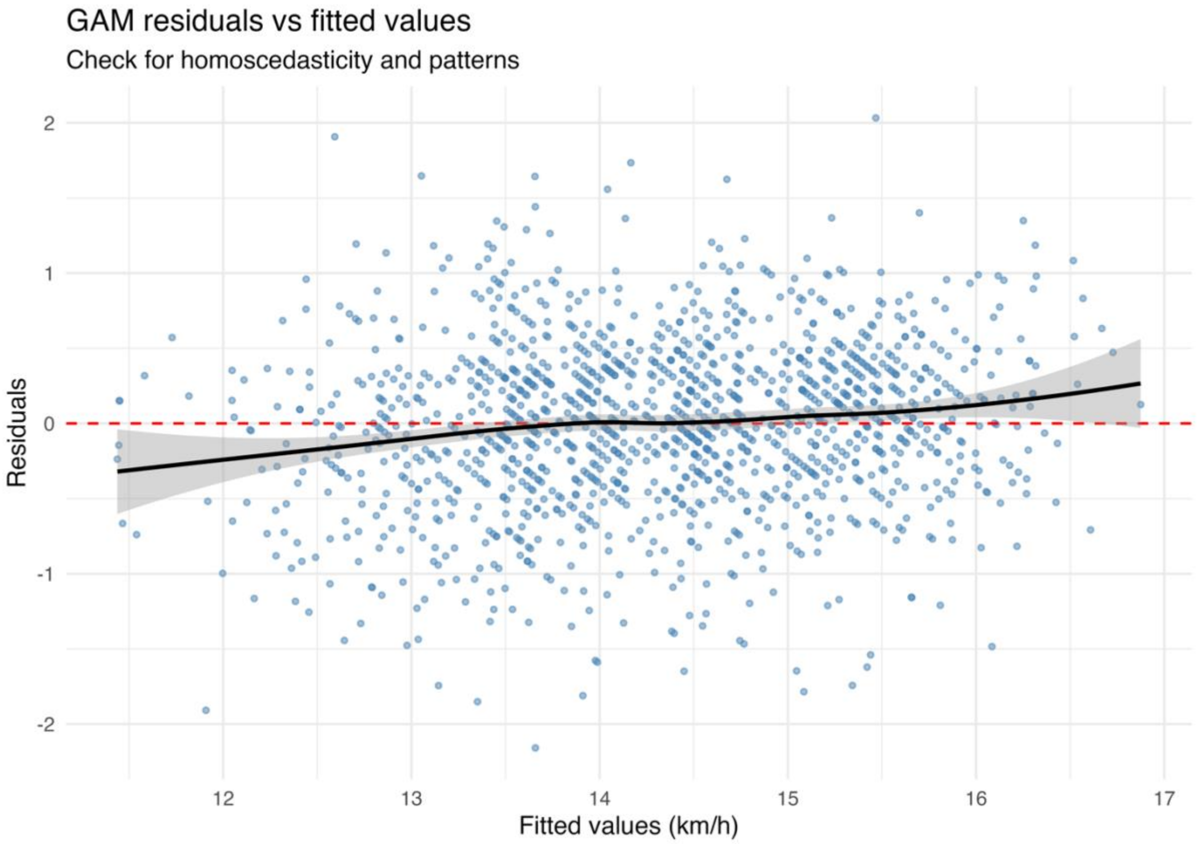

Supplementary 4. Figure 2. Residuals versus fitted values from the Generalized Additive Model (GAM).

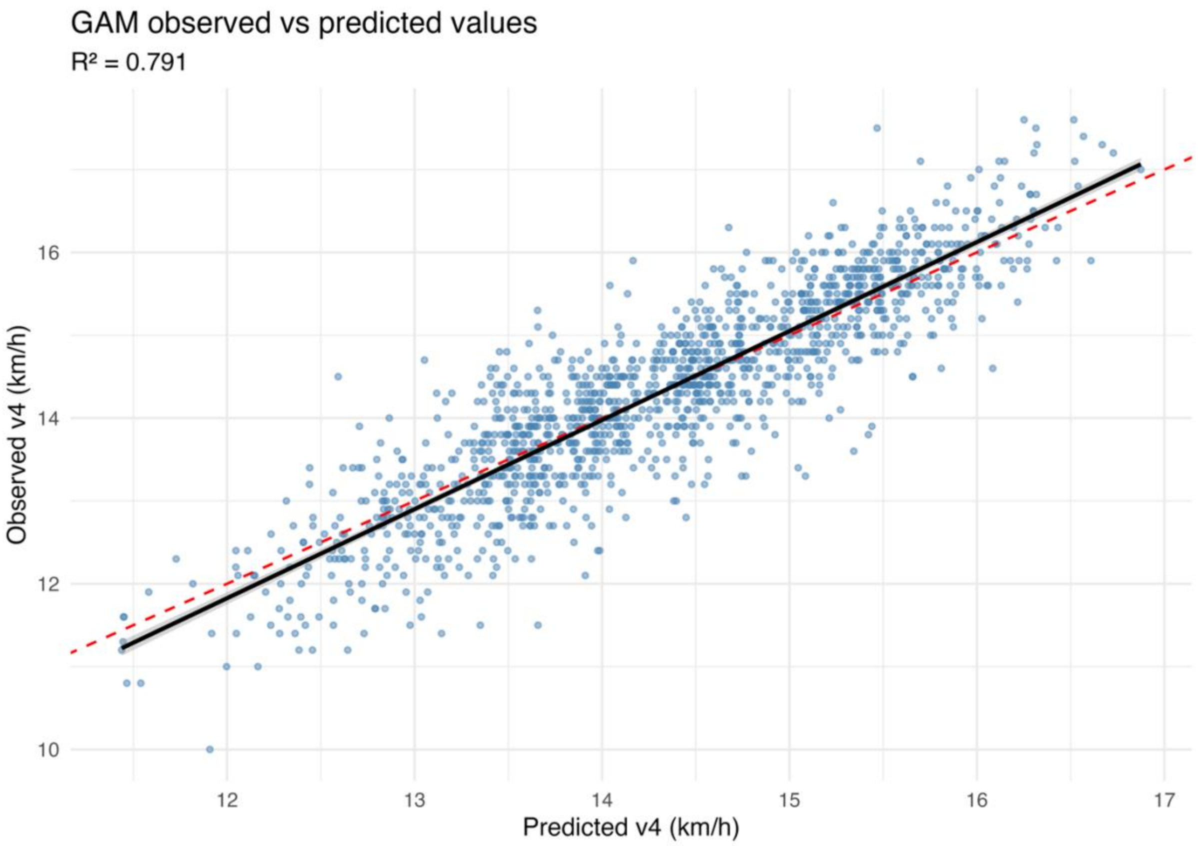

Supplementary 4. Figure 3. Observed versus predicted values from the Generalized Additive Model (GAM).

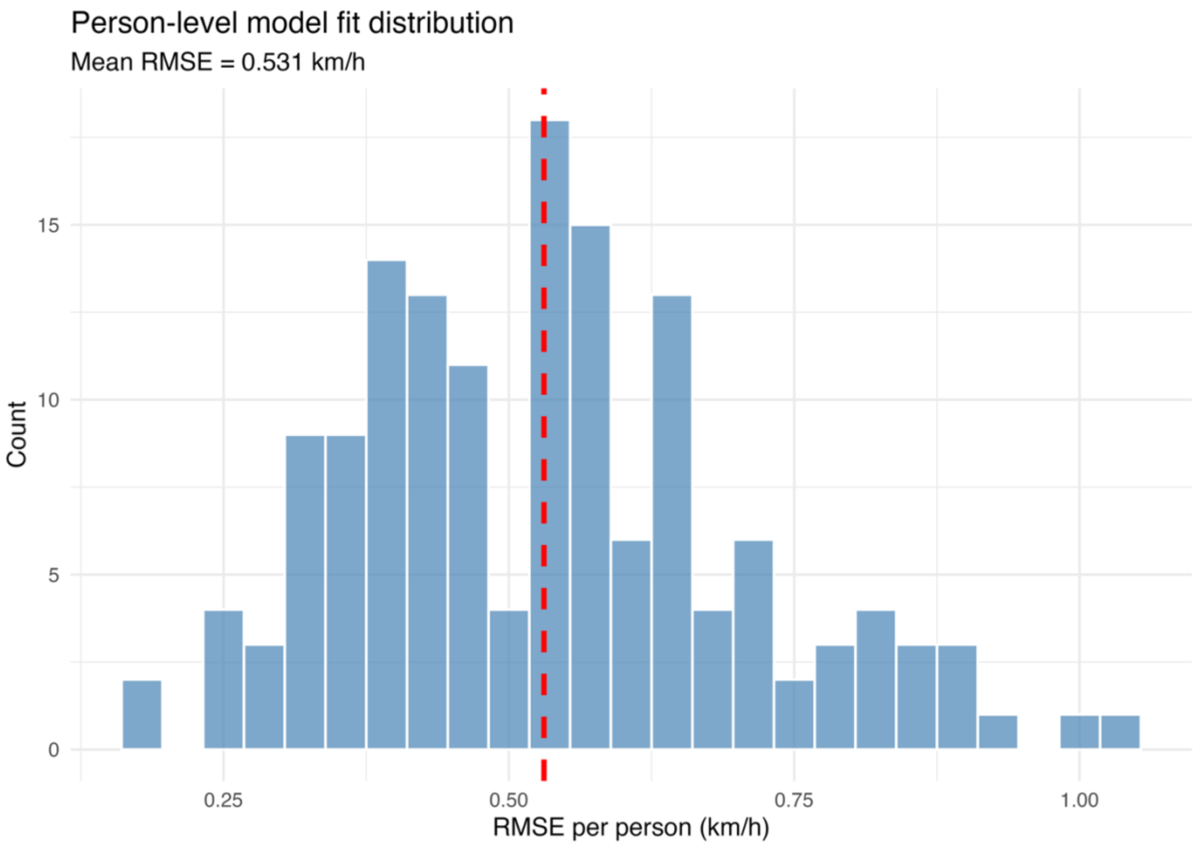

Supplementary 4. Figure 4. Distribution of player-level fit indices in the Generalized Additive Model (GAM).

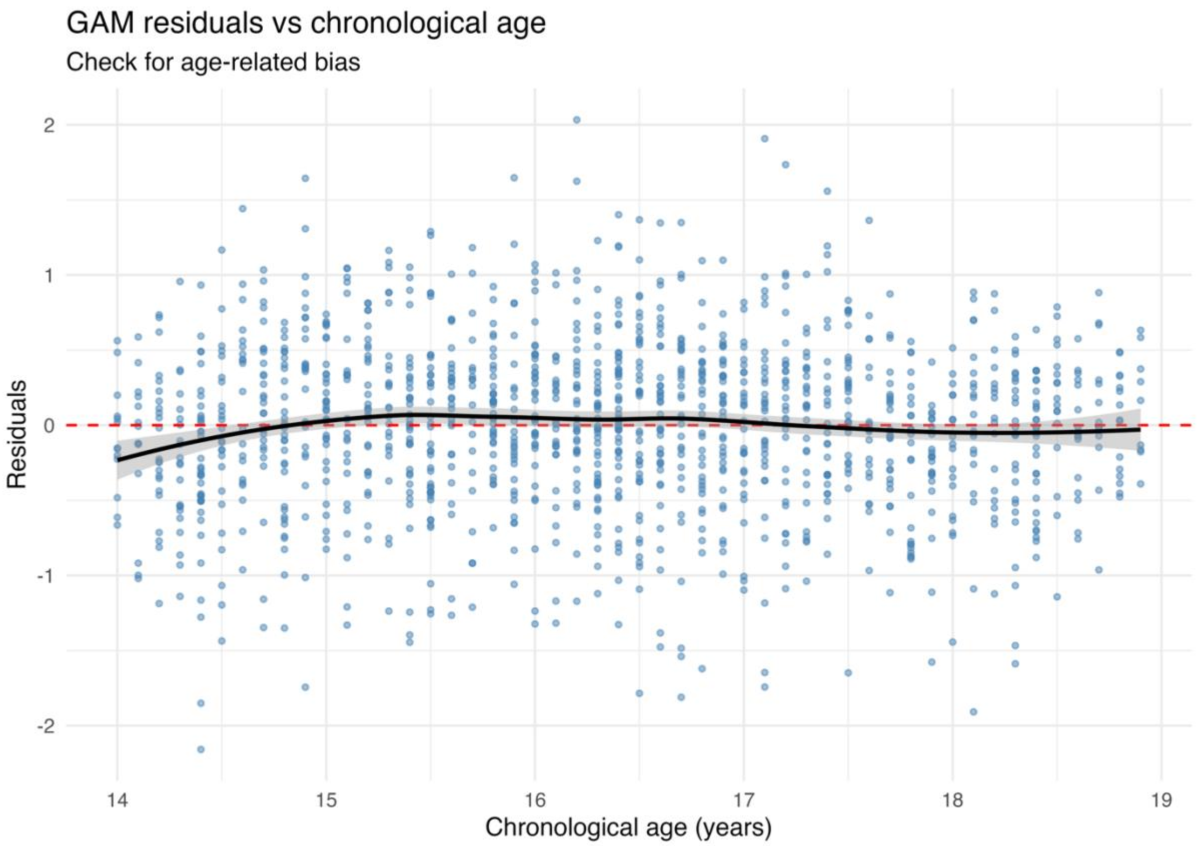

Supplementary 4. Figure 5. Residuals versus chronological age for the Generalized Additive Model (GAM).

<u>Step 2: Feature-Based clustering (Mclust diagnostics)</u>

Supplementary 4. Table 2. Mclust model selection results across cluster numbers and covariance structures.

| Number clusters | Model | BIC | AIC | loglik | Dunn | DB | CH | Silhouette |
|---|---|---|---|---|---|---|---|---|
| 6 | VEI | -4992 | 4647 | -2209 | 0.067 | 1.7 | 32.3 | 0.180 |
| 5 | VEI | -5144 | 4850 | -2327 | 0.067 | 1.7 | 32.7 | 0.166 |
| 4 | VEI | -5269 | 5026 | -2432 | 0.096 | 1.6 | 39.0 | 0.208 |
| 3 | VEI | -5570 | 5378 | -2625 | 0.086 | 1.9 | 28.4 | 0.148 |
| 3 | VVI | -5597 | 5321 | -2568 | 0.086 | 1.9 | 27.9 | 0.144 |
| 3 | EVI | -5765 | 5494 | -2657 | 0.062 | 1.7 | 37.4 | 0.208 |

Note: G refers to the number of clusters estimated in the model

Supplementary 4. Table 3: Final Mclust model quality metrics and interpretations.

| Metric | Value | Interpretation |
|---|---|---|
| Average Silhouette Width | 0.208 | Poor |
| Dunn | 0.787 | Good |
| Davies-Bouldin (DB) | 2.2 | Poor |
| Calinski-Harabasz (CH) | 15.4 | Higher is better |
| Bootstrap Stability (Mean ARI) | 0.61 | Good |
| Bootstrap Stability (SD) | 0.18 | - |
| Entropy | 0.02 | Good |
| Mean Uncertainty | 0.01 | Good |

464

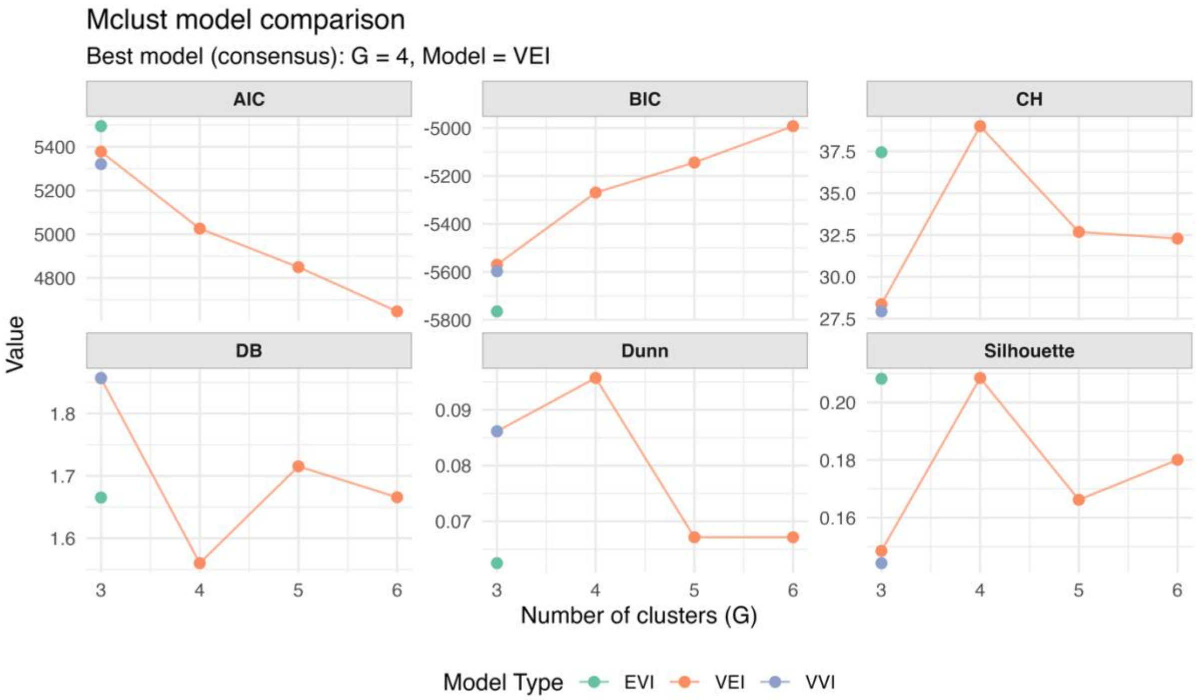

465

466 Supplementary 4. Figure 6. Comparison of Mclust model selection metrics across cluster numbers
467 and covariance structures. CH: Calinski-Harabasz; DB: Davies-Bouldin.

468

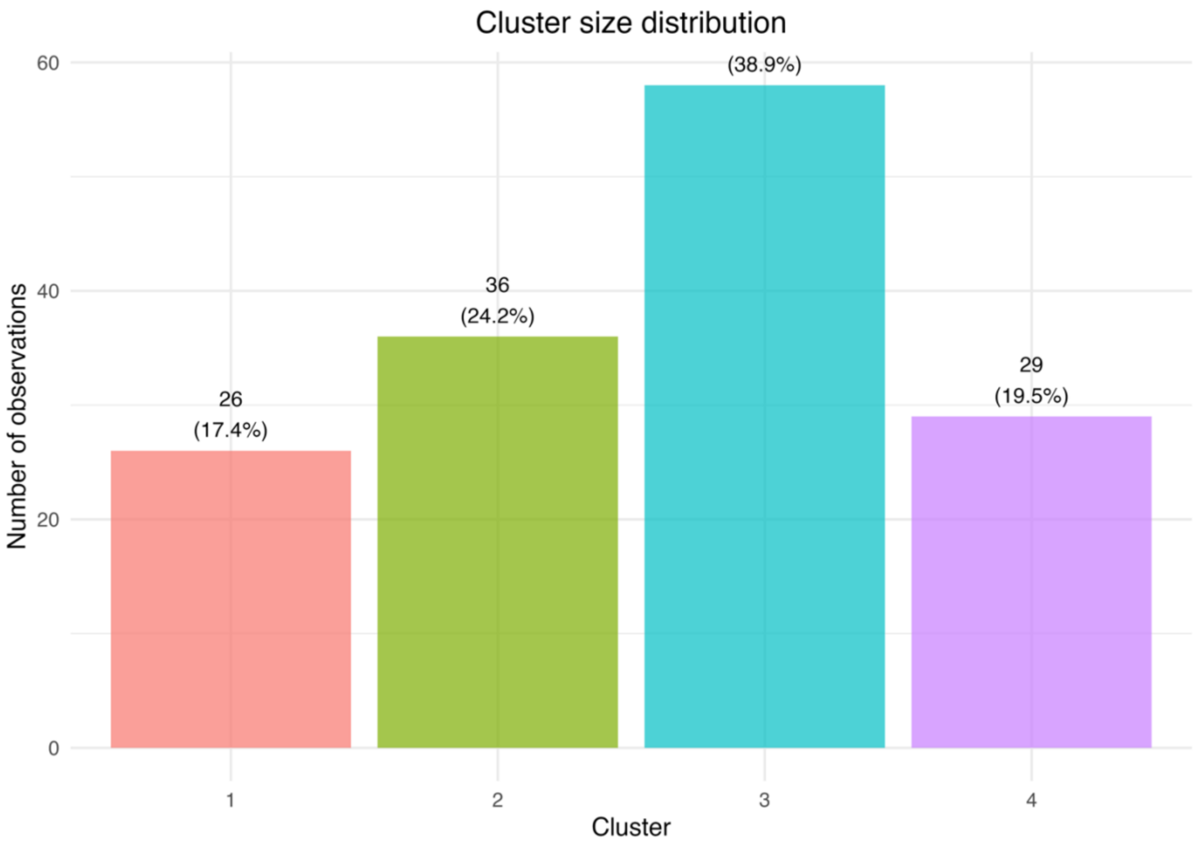

Supplementary 4. Figure 7. Cluster size distribution of the selected Mclust model.

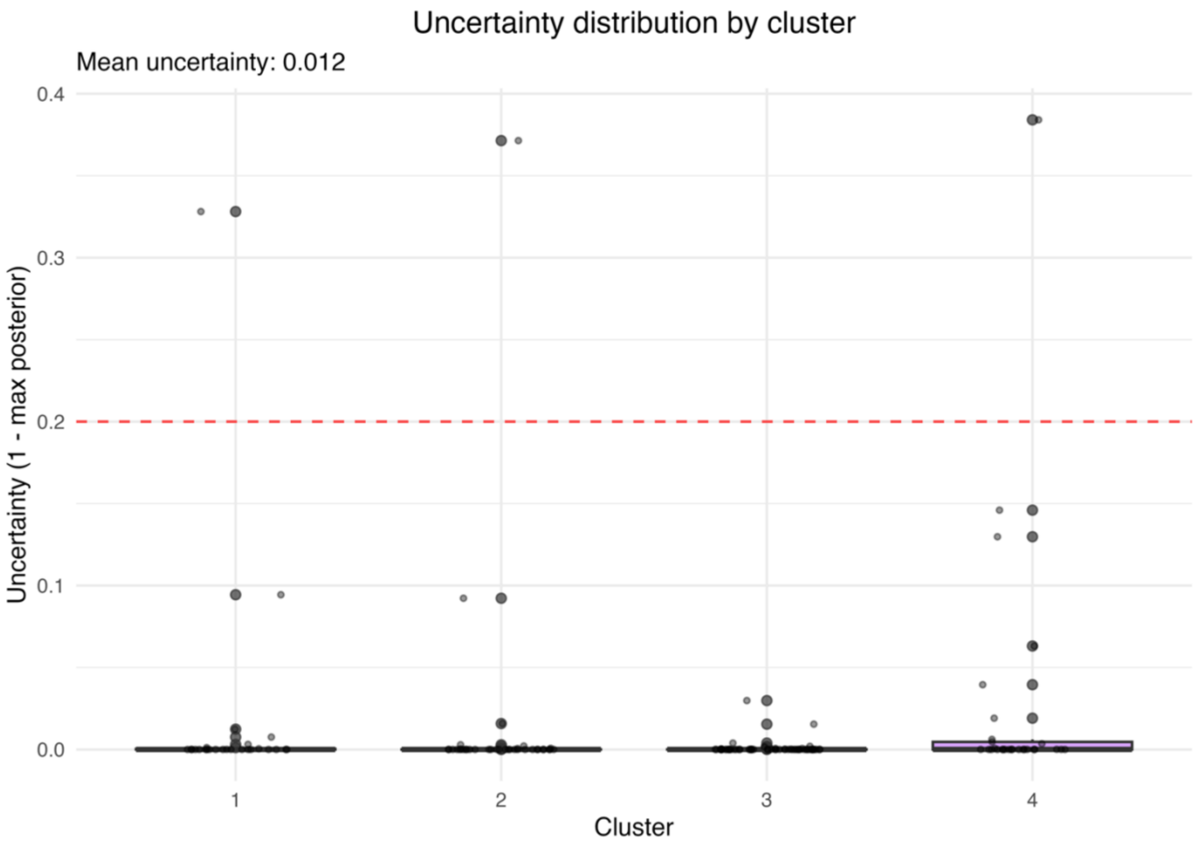

Supplementary 4. Figure 8. Distribution of classification uncertainty by cluster of the selected Mclust model.

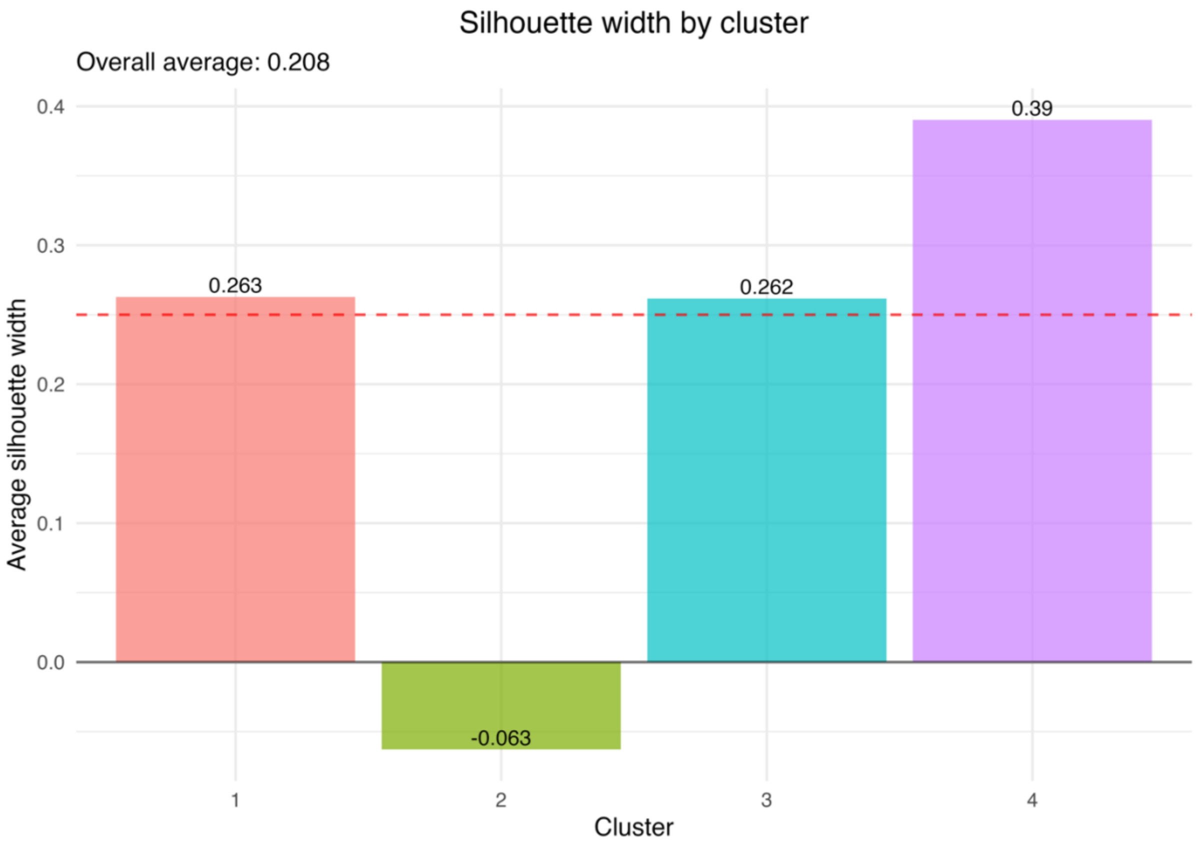

Supplementary 4. Figure 9. Silhouette width by cluster of the selected Mclust model.

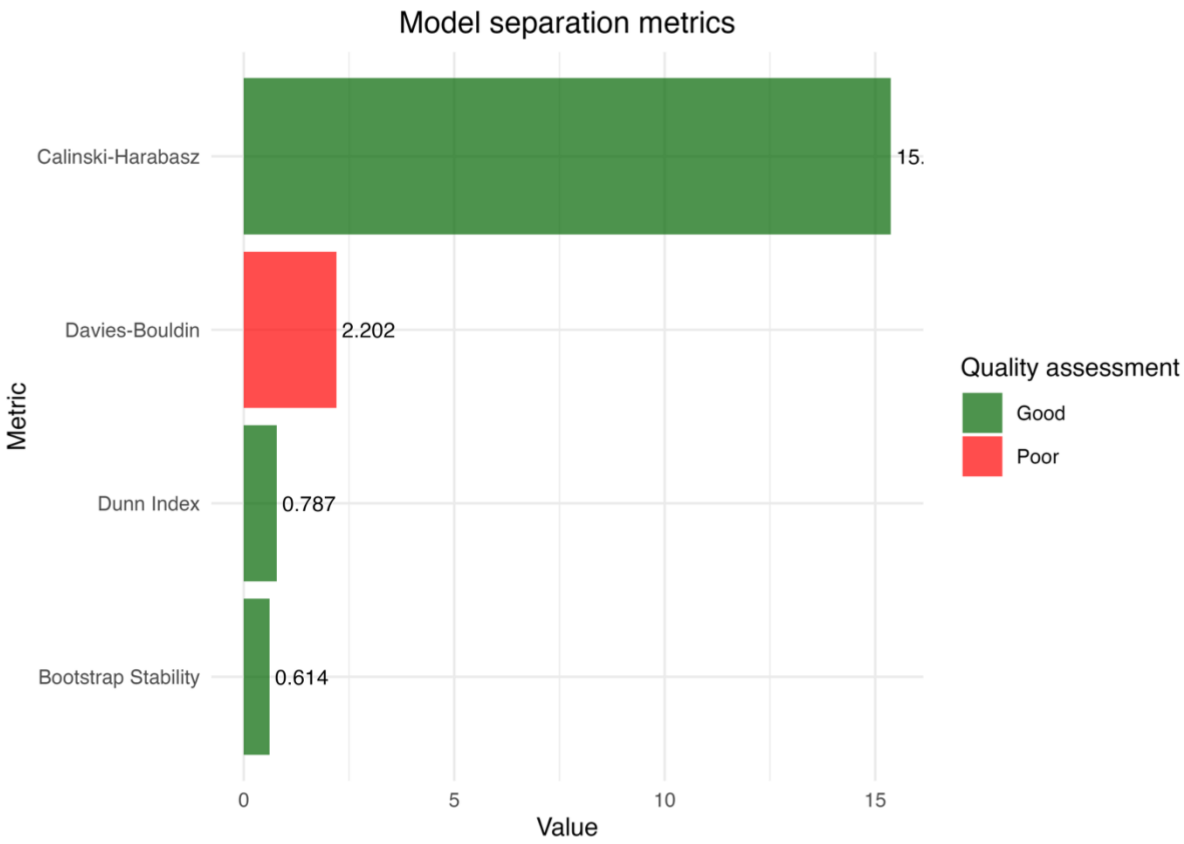

Supplementary 4. Figure 10. Comparison of separation metrics (Calinski-Harabasz, Davies-Bouldin, Dunn and Bootstrap stability) of the selected Mclust model.

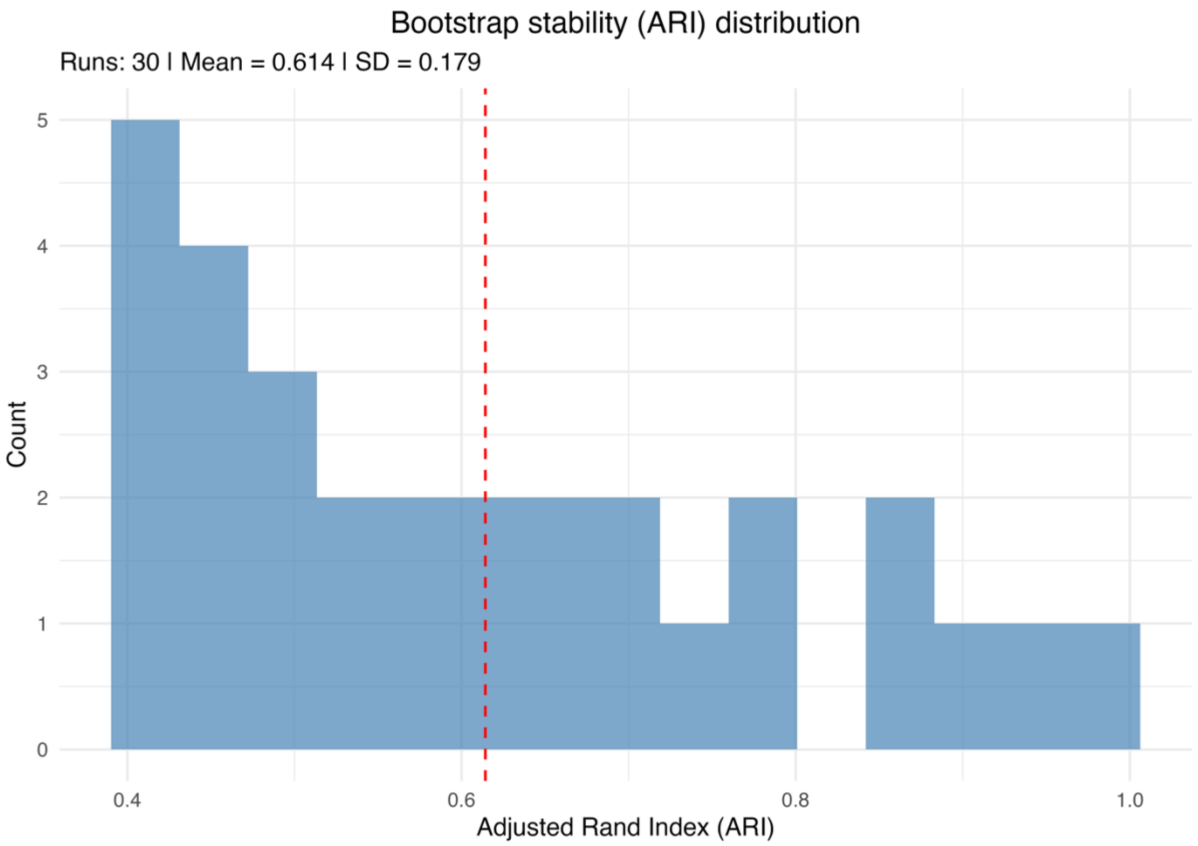

Supplementary 4. Figure 11. Bootstrap distribution of Adjusted Rand Index (ARI) values assessing cluster stability of the selected Mclust model.

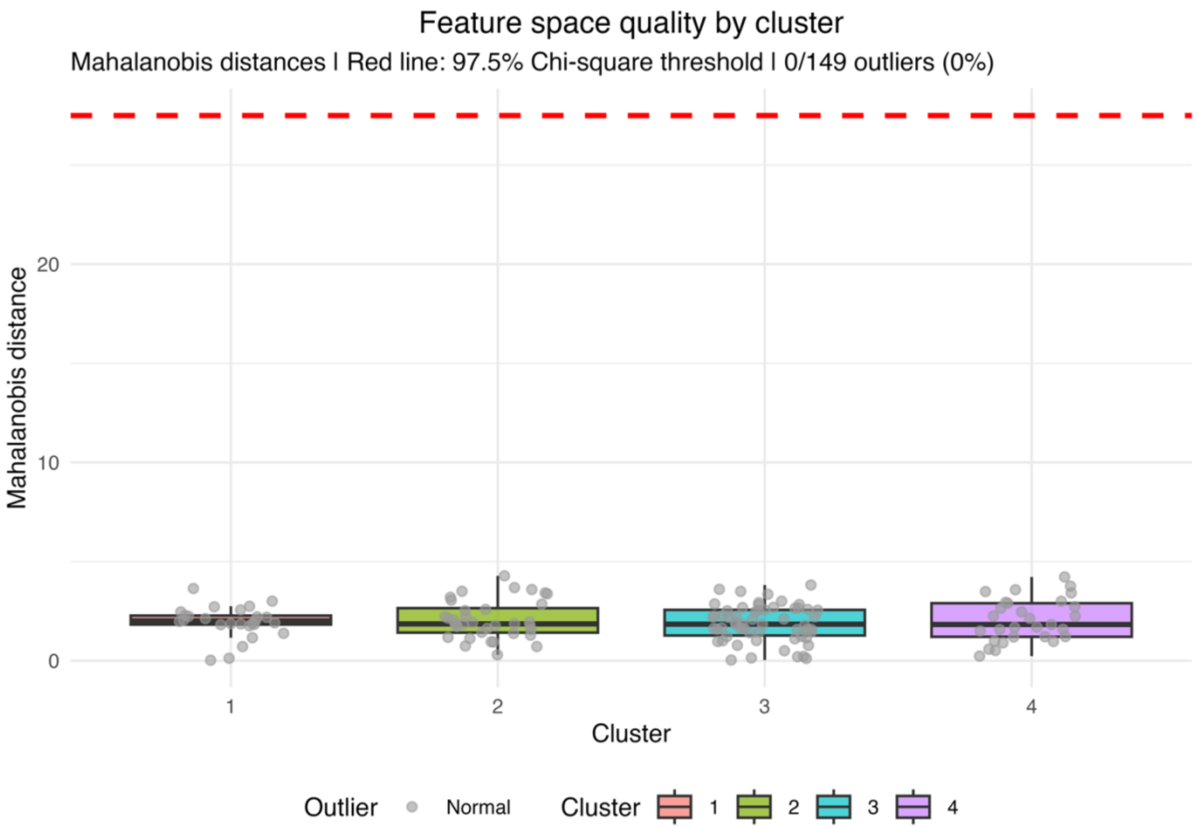

Supplementary 4. Figure 12. Mahalanobis distance by cluster representing feature space separation of the selected Mclust model.

***Model diagnostics for Latent Class Mixed Model (LCMM) approach***

<u>Latent Class Mixed Model (LCMM) selection diagnostics</u>

Supplementary 4. Table 4. Comprehensive Latent Class Mixed Model (LCMM) comparison across degrees of freedom (df) and cluster numbers (ng) for all computed models.

| **Model** | **df** | **ng** | **loglik** | **AIC** | **BIC** | **Dunn** | **Davies-Bouldin (DB)** | **Calinski Harabasz (CH)** | **Silhouette** | **converged** |
|---|---|---|---|---|---|---|---|---|---|---|
| df3_ng3 | 4 | 3 | -1599 | 3250 | 3328 | 0.005 | 3.1 | 6.1 | -0.022 | TRUE |
| df4_ng3 | 3 | 3 | -1607 | 3259 | 3328 | 0.004 | 3.0 | 7.8 | -0.118 | TRUE |
| df5_ng3 | 5 | 3 | -1594 | 3247 | 3334 | 0.005 | 5.6 | 4.2 | -0.042 | TRUE |
| df3_ng4 | 3 | 5 | -1582 | 3234 | 3339 | 0.005 | 3.4 | 17.2 | -0.062 | TRUE |
| df4_ng4 | 4 | 4 | -1588 | 3241 | 3340 | 0.005 | 4.8 | 6.6 | -0.077 | TRUE |
| df4_ng5 | 3 | 4 | -1603 | 3263 | 3350 | 0.004 | 7.2 | 6.9 | -0.136 | TRUE |
| df3_ng6 | 3 | 6 | -1577 | 3235 | 3358 | 0.005 | 5.1 | 16.8 | -0.027 | TRUE |
| df5_ng5 | 5 | 4 | -1594 | 3261 | 3372 | 0.004 | 11.5 | 7.1 | -0.145 | TRUE |

| | | | | | | | | | | |
|---|---|---|---|---|---|---|---|---|---|---|
| df3_ng5 | 4 | 6 | -1572 | 3238 | 3379 | 0.006 | 4.7 | 19.8 | -0.007 | TRUE |
| df5_ng4 | 5 | 5 | -1580 | 3251 | 3386 | 0.004 | 11.7 | 6.9 | -0.165 | TRUE |
| df4_ng6 | 4 | 5 | -1595 | 3270 | 3390 | 0.004 | 5.8 | 5.8 | -0.157 | TRUE |
| df5_ng6 | 5 | 6 | -1568 | 3242 | 3401 | 0.006 | 5.0 | 21.6 | 0.001 | TRUE |

Note: df refers to the degrees of freedom of the natural spline used to model the trajectory shape, and ng refers to the number of latent classes (clusters) estimated in the model.

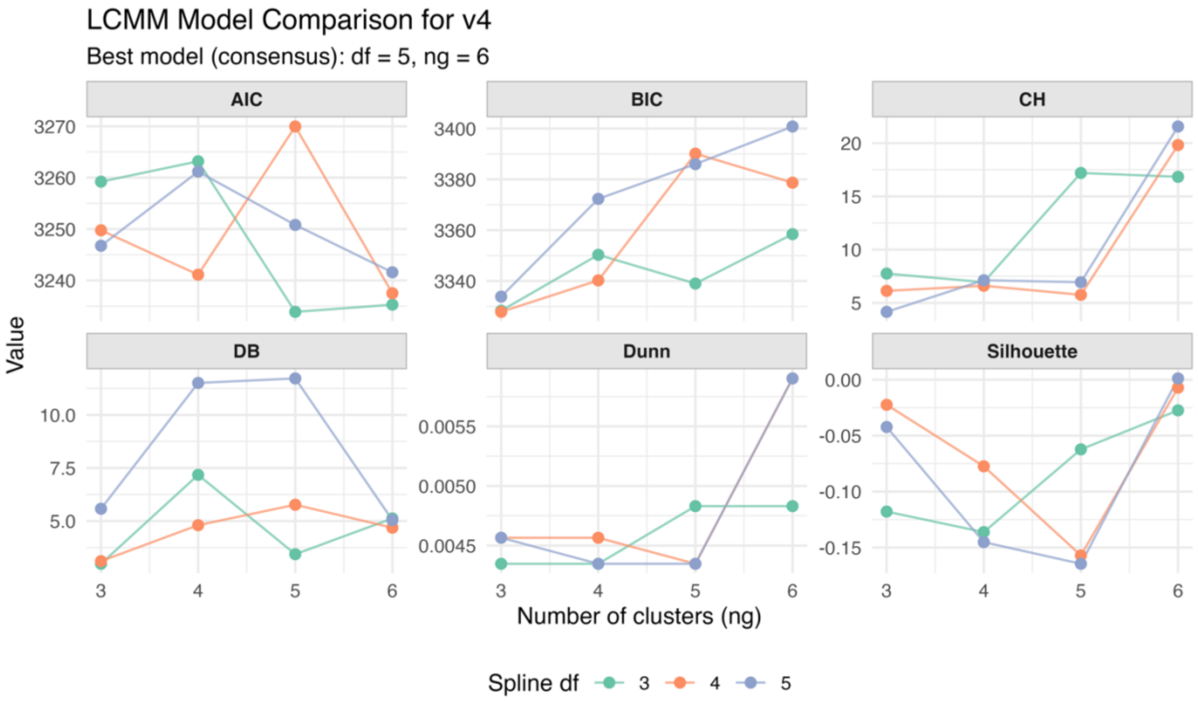

Supplementary 4. Figure 13. Comparison of Latent Class Mixed Model (LCMM) selection metrics across degrees of freedom (df) and cluster numbers (ng). CH: Calinski-Harabasz; DB: Davies-Bouldin

<u>Latent Class Mixed Model (LCMM) clustering quality diagnostics</u>

Supplementary 4. Table 5. Final Latent Class Mixed Model (LCMM) clustering quality metrics and interpretations.

| Metric | Value | Interpretation |
|---|---|---|
| Average Silhouette Width | 0.044 | Poor |
| Dunn | 0.841 | Good |
| Davies-Bouldin (DB) | 7.1 | Poor |
| Calinski-Harabasz (CH) | 8.7 | Higher is better |

| | | |
|---|---|---|
| Bootstrap Stability (Mean ARI) | 0.41 | Moderate |
| Bootstrap Stability (SD) | 0.14 | - |
| Entropy | 0.26 | Good |
| Mean Uncertainty | 0.18 | Good |

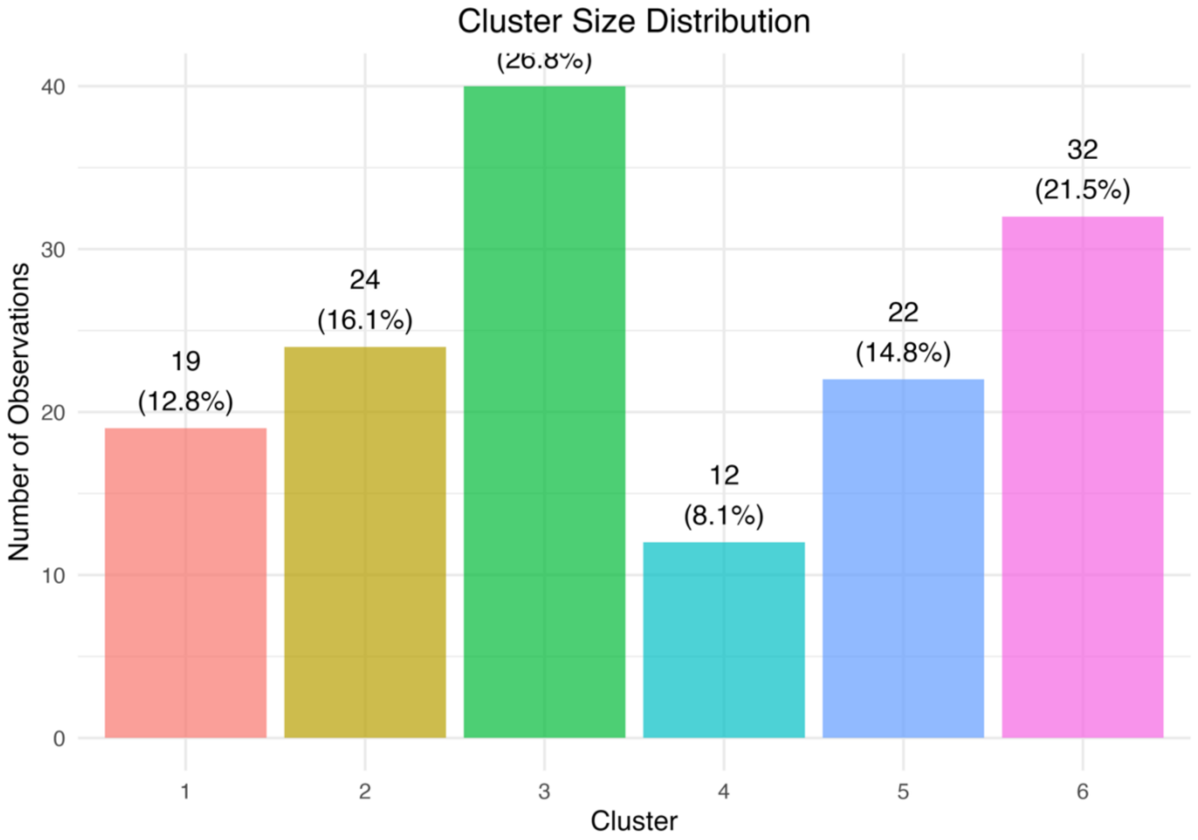

Supplementary 4. Figure 14. Cluster size distribution of the selected Latent Class Mixed Model (LCMM).

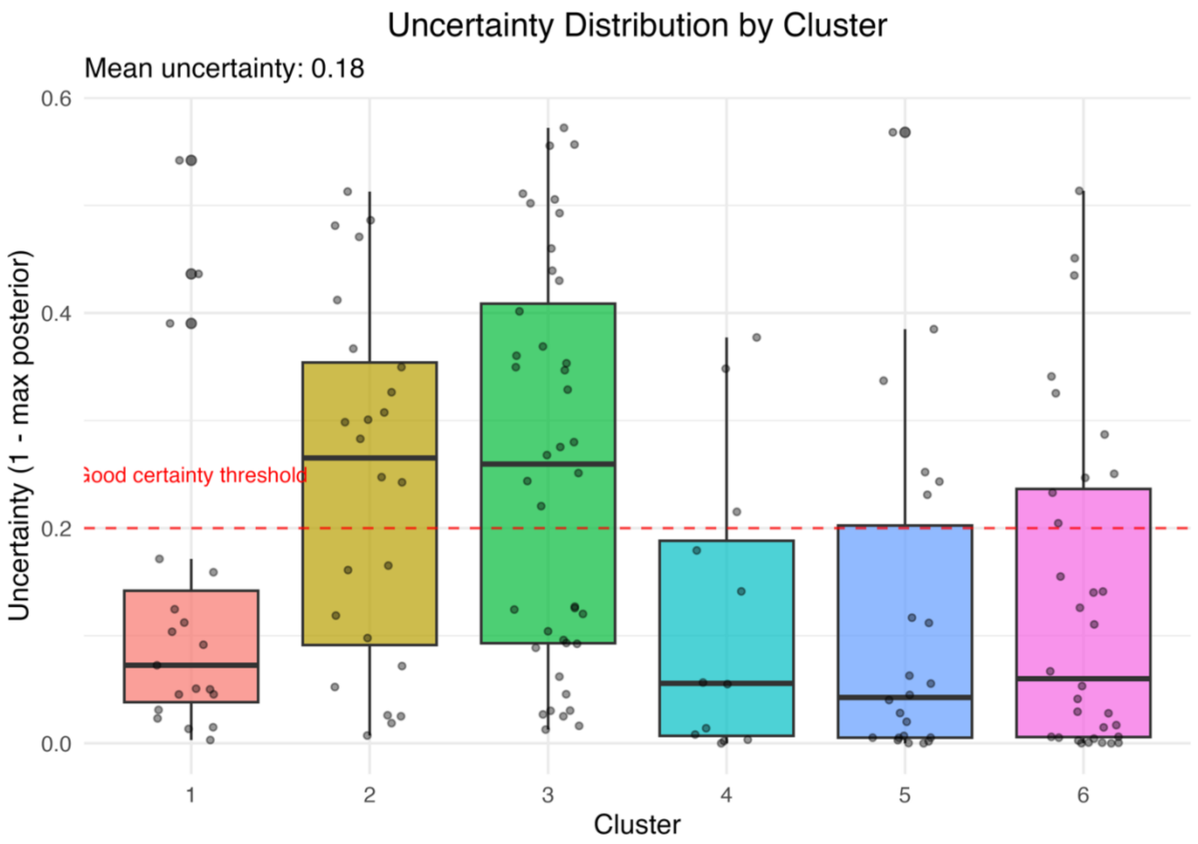

511

512 Supplementary 4. Figure 15. Distribution of classification uncertainty by cluster of the selected Latent
513 Class Mixed Model (LCMM).

514

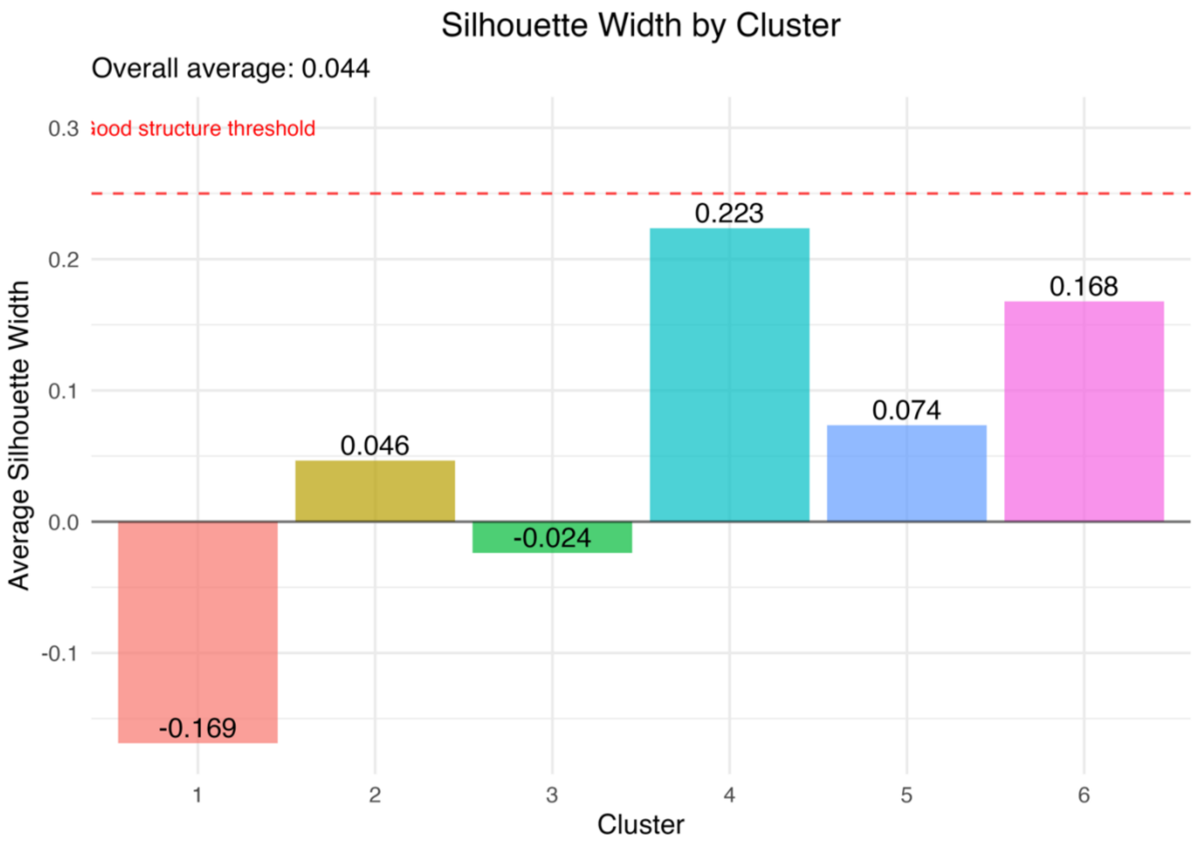

Supplementary 4. Figure 16. Silhouette width by cluster of the selected Latent Class Mixed Model (LCMM).

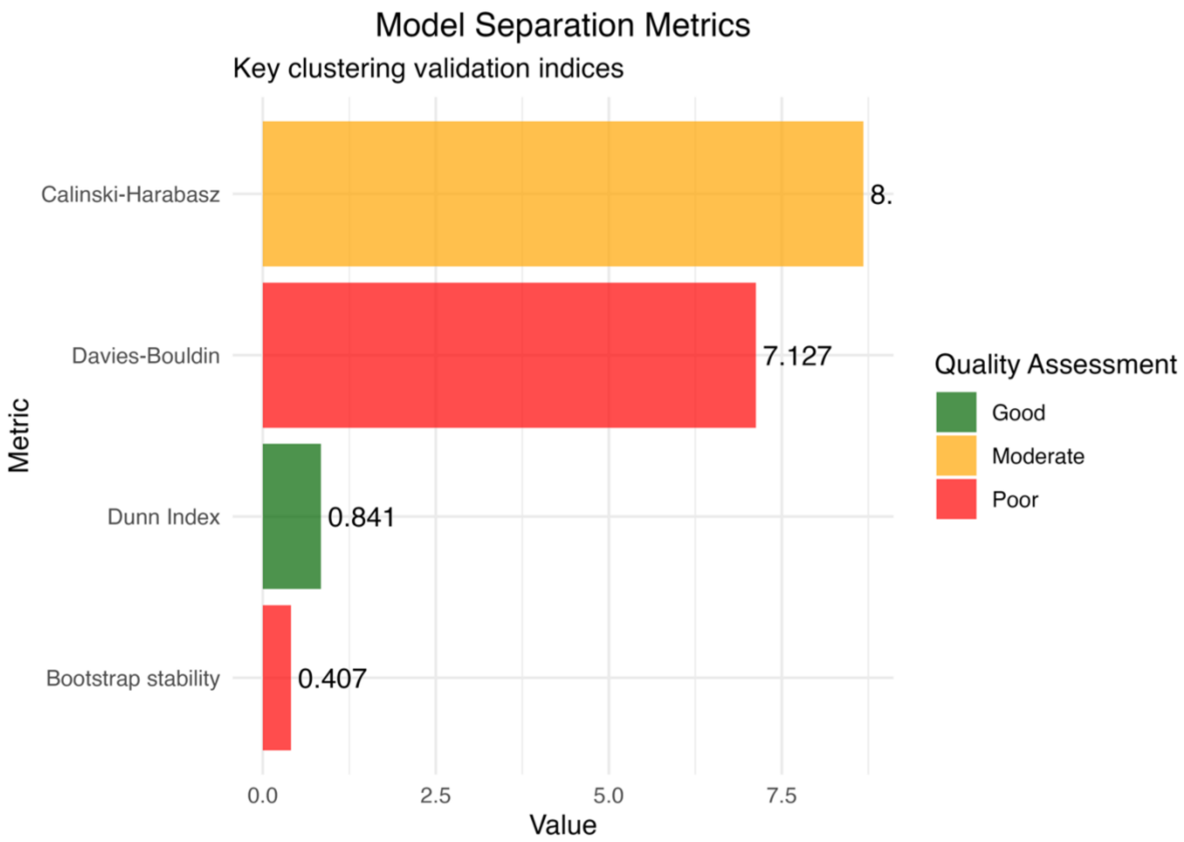

Supplementary 4. Figure 17. Comparison of separation metrics (Calinski-Harabasz, Davies-Bouldin, Dunn and Bootstrap stability) of the selected Latent Class Mixed Model (LCMM).

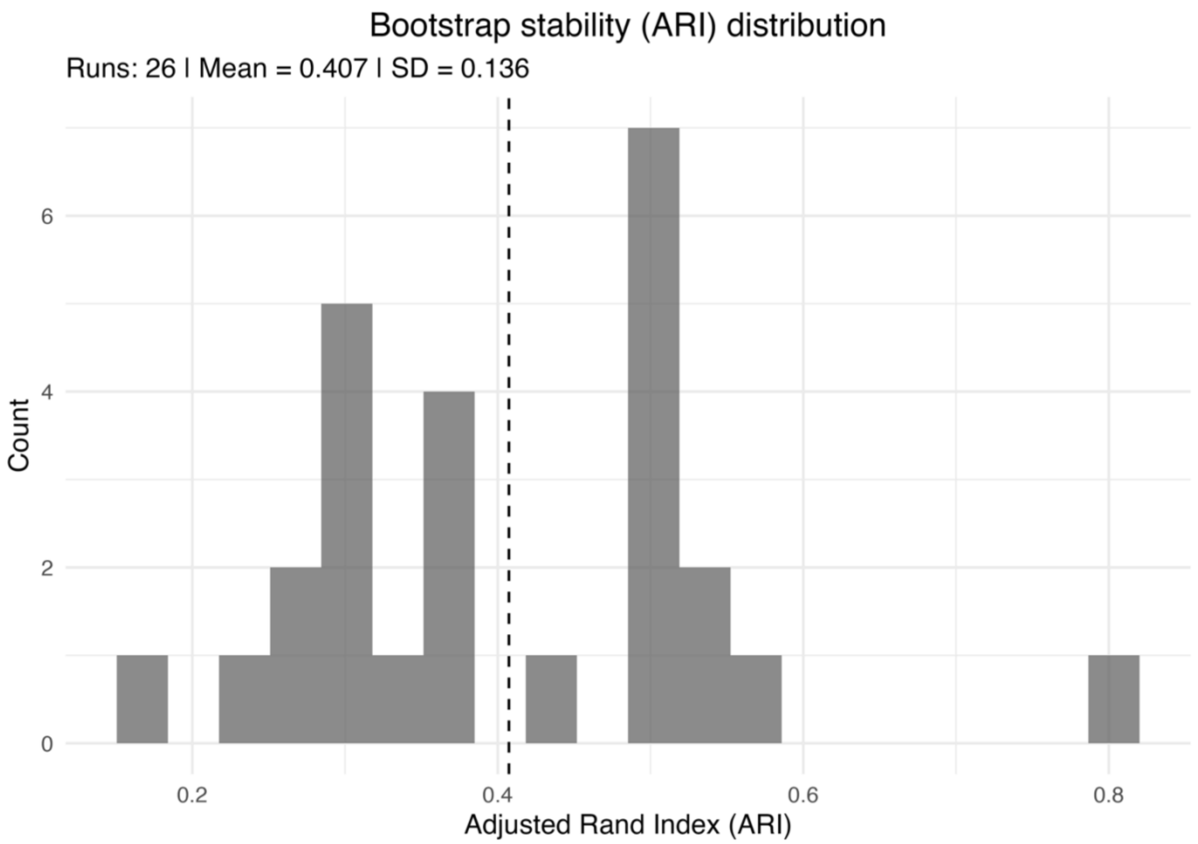

Supplementary 4. Figure 18. Bootstrap distribution of Adjusted Rand Index (ARI) values assessing cluster stability of the selected Latent Class Mixed Model (LCMM).

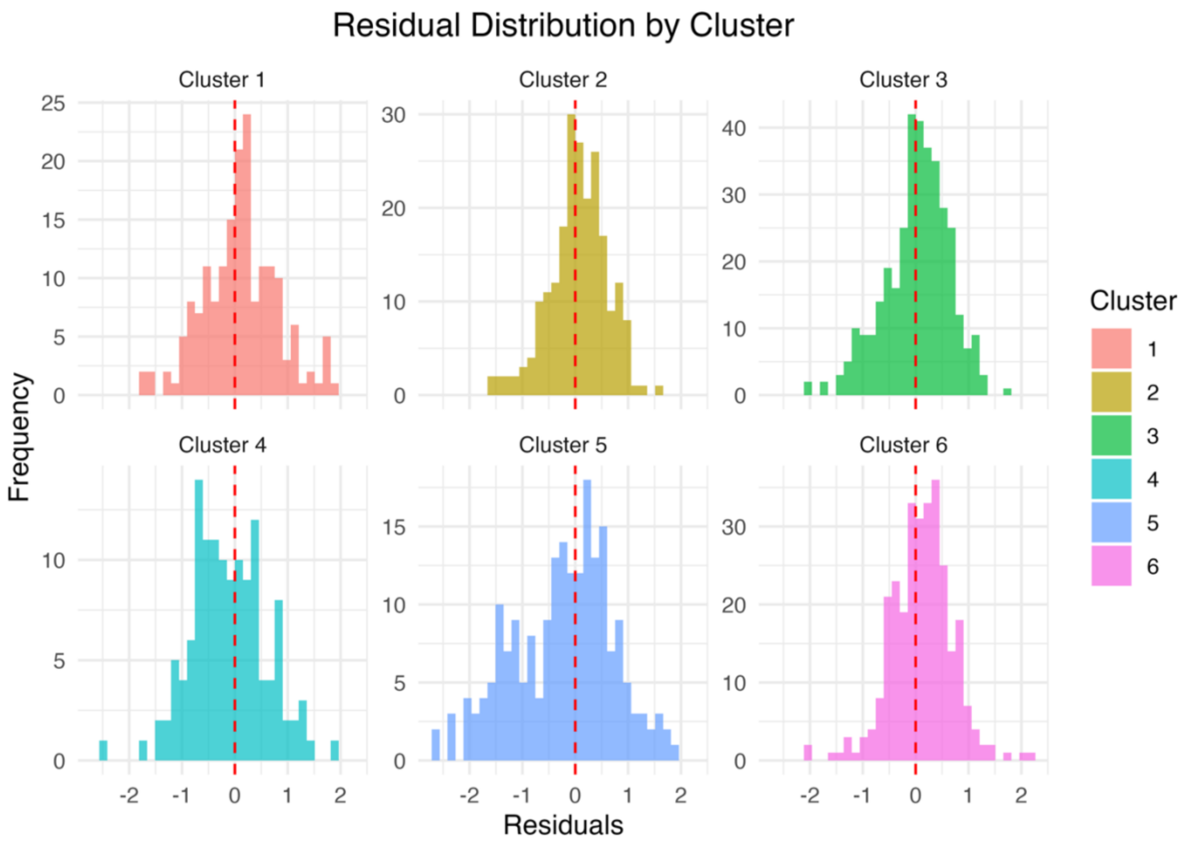

Supplementary 4. Figure 19. Distribution of residuals by cluster from the selected Latent Class Mixed Model (LCMM).

### Supplementary 5. Comparative quality assessment across both outcome measures

Supplementary 4. Table 6. Side-by-side comparison of clustering quality metrics between the Hybrid (GAM + Mclust) and Latent Class Mixed Model (LCMM) approaches for vIANT and v4.

| Outcome measure | Approach | Number clusters | BIC | AIC | Silhouette | Dunn | Calinski Harabasz (CH) | Bootstrap Stability (Mean ARI) | Entropy | Mean Uncertainty |
|---|---|---|---|---|---|---|---|---|---|---|
| vIANT | Hybrid | 4 | -5073 | 4830 | 0.207 | 0.880 | 15.1 | 0.59 | 0.02 | 0.01 |
| vIANT | LCMM | 6 | 2581 | 2422 | -0.002 | 0.831 | 7.8 | 0.22 | 0.26 | 0.19 |
| v4 | Hybrid | 4 | -5269 | 5026 | 0.208 | 0.787 | 15.4 | 0.61 | 0.02 | 0.01 |
| v4 | LCMM | 6 | 3401 | 3242 | 0.044 | 0.841 | 8.7 | 0.41 | 0.26 | 0.18 |

*Brief interpretation*

The comparative assessment reveals substantial differences in clustering quality between the two methodological approaches across both outcome measures. The Hybrid approach (GAM + Mclust) demonstrated consistently superior performance compared to the Latent Class Mixed Model (LCMM) approach across multiple validation criteria. Specifically, the Hybrid approach achieved markedly higher silhouette coefficients (vIANT: 0.207 vs -0.002; v4: 0.208 vs 0.044), indicating substantially better cluster separation and more confident individual assignments. Bootstrap stability analysis further supported the robustness of the Hybrid approach, with mean Adjusted Rand Index values approximately 2.5-fold higher for vIANT (0.59 vs 0.22) and 1.5-fold higher for v4 (0.61 vs 0.41), demonstrating greater consistency in cluster assignments across resampled datasets. The Hybrid approach also exhibited lower entropy statistics (0.02 vs 0.26 for both outcomes), reflecting clearer cluster boundaries with minimal assignment ambiguity.

While both approaches identified distinct developmental trajectory patterns, the Latent Class Mixed Model (LCMM) approach consistently selected solutions with more clusters (6 vs 4) alongside weaker separation metrics, suggesting potential overfitting or identification of less substantive subgroups. The negative silhouette coefficient observed for vIANT in the LCMM approach (-0.002) indicates that individuals were, on average, positioned closer to neighboring clusters than to their assigned cluster, raising concerns about the meaningfulness of the identified groupings. Model fit criteria (BIC and AIC) differed substantially between approaches due to fundamental differences in modeling frameworks, precluding direct comparison. However, the convergence of multiple independent validation metrics (including silhouette width, bootstrap stability, and entropy) consistently favored the Hybrid approach (GAM + Mclust), supporting its selection as the primary analytical framework for identifying distinct endurance development trajectories in youth soccer players.

### Supplementary 6. Overall mean trajectory generalized additive model information

#### *Model Information*

*Model Specification*

The overall population mean trajectory was estimated using generalized additive models (GAMs) with the following specification:

Outcome ~ s(CalendarAge, bs = [basis], k = [k]) + s(PersonNumericId, bs = "re")

where the first smooth term captures the non-linear age effect and the second term represents random intercepts for players to account for repeated measures.

*Model Selection Procedure*

Optimal model parameters were determined through systematic evaluation of:

Basis functions: Thin plate splines (tp), cubic regression splines (cr), shrinkage cubic splines (cs), and P-splines (ps)

Smoothness parameters: k = 3 to 6

Model selection was based on minimizing the Akaike Information Criterion (AIC). All models were fitted using restricted maximum likelihood (REML) estimation.

#### *Results*

Summary of the model specifications using 149 players and 1380 observations for the two outcome measures lactate concentration at the individual aerobic threshold (vIANT) and velocity at the absolute lactate concentration of 4 mmol.L$^{-1}$ (v4).

| Outcome measure | Best basis function | Optimal k value | AIC | Deviance explained | R-squared |
|---|---|---|---|---|---|
| vIANT | tp | 6 | 2158 | 76.0% | 0.732 |
| v4 | ps | 4 | 2991 | 72.0% | 0.687 |

The adequacy of the selected basis dimensions was evaluated using the k.check() function, which tests whether the chosen k value provides sufficient flexibility to capture the underlying relationship.

| *Lactate concentration at the individual aerobic threshold (vIANT)* | | | | |
|---|---|---|---|---|
| **Smooth term** | **k'** | **Effective degrees of freedom (edf)** | **k-index** | **p-value** |
| s(CalendarAge) | 5 | 3.6 | 1.05 | 0.985 |
| s(PersonNumericId) | 149 | 141.6 | na | Na |
| *Absolute lactate concentration of 4 $mmol.L^{-1}$ (v4)* | | | | |
| s(CalendarAge) | 3 | 2.7 | 1.01 | 0.6425 |
| s(PersonNumericId) | 149 | 140.2 | na | na |

For both models, the k-index values are greater than 1 (vIANT: 1.05; v4: 1.01) and p-values are non-significant ($p > 0.05$), indicating that the selected k values provide adequate basis dimensions. The effective degrees of freedom (edf) for the age smooth terms (vIANT: 3.6; v4: 2.7) are substantially lower than the maximum possible (k' = 5 and 3, respectively), demonstrating that the smoothing penalty is appropriately constraining model complexity without underfitting. The high edf values for the player random effects (vIANT: 141.6; v4: 140.2 out of 149) reflect substantial between-player heterogeneity in baseline performance levels, justifying the inclusion of random intercepts in the model specification.

*Prediction*

Population-level mean trajectories were obtained by generating predictions across the age range (14–19 years, 0.1-year increments) while excluding the random effect term from predictions. This approach provides marginal mean estimates with appropriate standard errors reflecting uncertainty in the fixed effect only.

***Overall interpretation***

The high deviance explained (72–76%) and R-squared values (0.69–0.73) indicate that the models capture the majority of systematic variation in endurance performance development. The random intercepts account for substantial between-player heterogeneity, while the smooth age terms characterize the average developmental trajectory across the sample.
